# Cost-effectiveness of risk-stratified screening for cervical cancer in cohorts vaccinated against human papillomavirus with moderate vaccination coverage

**DOI:** 10.1101/2025.06.12.25329488

**Authors:** Tiago M. de Carvalho, Johannes Berkhof, Johannes A. Bogaards

**Affiliations:** Amsterdam UMC, location Vrije Universiteit Amsterdam, Dept. Epidemiology and Data Science, De Boelelaan 1117, Amsterdam, The Netherlands; Amsterdam Public Health, Amsterdam, The Netherlands; Cancer Center Amsterdam, Amsterdam, The Netherlands; Amsterdam Institute for Immunology and Infectious Diseases, Amsterdam, The Netherlands

## Abstract

**Background:** Cervical cancer screening in the Netherlands consists of human papillomavirus (HPV) testing followed by cytological triage at age 30, 35, 40, 50 and 60 years. Women are also invited at age 45, 55 and 65 years if they did not test HPV-negative five years earlier (risk-based invitation). With influx of birth cohorts vaccinated against HPV, de-intensification may be needed to maintain a cost-effective program.

**Methods:** We used an updated and recalibrated model of type-specific HPV transmission and cervical carcinogenesis to estimate the cost-effectiveness of 16 strategies with reduced screening intensity. Strategies varied by starting age, screening interval, and number of risk-based invitations, possibly stratified for HPV vaccination status. Cost-effectiveness was measured by net monetary benefit (NMB). A positive NMB indicates that a strategy is cost-effective compared to the current policy.

**Findings:** Two strategies without stratification for HPV vaccination status had a positive NMB. An NMB of €1.6 million per 100,000 women was obtained when, compared to the current strategy, the invitation at age 35 was based on the HPV-test result at age 30. An NMB of €1.8 million per 100,000 women was obtained when women were invited at age 30 and subsequent invitations from age 35 to 65 were all risk-based. If reduced screening was only applied to vaccinated women, the highest NMB was €1.3 million per 100,000 women.

**Interpretation:** Reducing screening in HPV-vaccinated cohorts is cost-effective when re-inviting women after ten years if they test HPV-negative. Stratification for HPV vaccination status does not improve the cost-effectiveness of screening.

## INTRODUCTION

Screening has been the main pillar of cervical cancer prevention for several decades. The recognition that cervical cancer is caused by sexually transmitted oncogenic types of human papillomavirus (HPV) has led to the switch from a cytology-based screening program to primary high-risk HPV (hrHPV) testing with cytological triage after a positive test result [1]. In the Netherlands, HPV-based screening is currently performed at five fixed screening rounds at ages 30, 35, 40, 50 and 60 years. Additionally, risk-based screening rounds are implemented at ages 45, 55, and 65 years, meaning that an invitation is sent to all women who had a positive hrHPV-test or did not attend screening five years earlier.

Routine HPV vaccination for 12-year-old girls started in the Netherlands in 2010, using the bivalent vaccine targeting HPV-16 and HPV-18. Additionally, in 2009, girls between ages 14 and 17 (birth cohorts 1993-1996) were vaccinated during a one-off catch-up campaign [2]. Since 2023, birth cohorts of HPV-vaccinated women reached age 30 and entered the Dutch cervical cancer screening program. HPV vaccination lowers the risk of cervical cancer and its precursors [3], and unvaccinated women benefit indirectly due to herd effects [4]. The influx of HPV-vaccinated birth cohorts into cervical cancer screening programs will likely alter the balance between the benefits and harms of screening, due to the reduction in cervical cancer risk [5–7]. Hence, de-intensification of screening may be needed to maintain a cost-effective program.

It is not obvious that the current screening intensity can be reduced in HPV-vaccinated cohorts in the Netherlands, given that the Dutch screening program already is one of the least intensive screening programs in Europe. Additionally, the coverage of HPV vaccination may be too low to provide sufficient indirect protection to unvaccinated women and de-intensification of the screening program could lead to an increased cervical cancer risk for unvaccinated women. In this context, the Netherlands represents an interesting case study because it has a moderate vaccination coverage of around 60%, similar to Germany and Italy, but substantially lower than Sweden and the UK [8].

Given the expected reduction in cervical cancer incidence due to HPV vaccination in the Dutch population, a re-evaluation of the benefits and harms of the current Dutch screening program is warranted. The intensity of the screening program can be reduced in several ways, by starting screening later, by extending the screening interval or by expanding risk-based invitations based on the HPV-test result in the previous round. De-intensification could be limited to vaccinated women only, to ensure that cancer risk does not increase for unvaccinated women. Such a strategy would require data linkage between the vaccine registry and the cancer screening organization, and may be difficult to implement because of logistical challenges and privacy reasons. Therefore, the cost-effectiveness of stratification based on HPV vaccination status requires careful consideration.

In this study we evaluate less intensive screening strategies for the first ten HPV-vaccinated birth cohorts (1993-2002) in the Netherlands, who will enter the Dutch screening program between 2023 and 2032. We perform a cost-effectiveness analysis of these cohorts based on hybrid model for type-specific HPV transmission and cervical cancer progression [9,10], informed by Dutch demographics, sexual surveys and recent epidemiological data on HPV prevalence, HPV-based screening outcomes and cervical cancer incidence.

## METHODS

### Modelling overview

To estimate the health and economic effects of cervical cancer screening in a partly vaccinated population, we combined a deterministic HPV transmission model and a microsimulation model of cervical cancer progression.

The HPV transmission model describes the transmission of 14 hrHPV infection types (HPV-16, - 18, -31, -33, -35, -39, -45, -51, -52, -56, -58, -59, -66, -68). HPV transmission and the natural history of type-specific infection are described by a set of partial differential equations in a compartmental SIRS (Susceptible, Infected, Recovered/Immune, Susceptible) model [9]. The main output of the transmission model for use in the cancer progression model concerns type-, age- and cohort-specific HPV infection incidences, stratified by categories of sexual activity in the heterosexual population [10].

The microsimulation model of cervical carcinogenesis describes the joint natural history of multiple-type HPV infections to cervical cancer and death (Supplementary Figure S.1). Type-specific HPV infection incidences are informed by the HPV transmission model. Once acquired, an HPV infection may clear or develop into a high-grade precancerous lesion (i.e. CIN2/3: cervical intraepithelial neoplasia grade 2 or 3). To ensure consistency, the natural history of type-specific HPV infection to CIN2/3 caused by the corresponding infection type employs common model parameters for the HPV transmission model and the cervical cancer progression model. Natural history parameters describing progression from infection up to CIN2/3 were estimated in previous publications based on longitudinal follow-up in the POBASCAM study (Supplementary Appendix A.1) [9–11]. Furthermore, durations from non-regressive CIN2/3 lesions to invasive cancer were previously estimated by statistically linking the age distributions of CIN2/3 and cervical cancer incidence, based on data from the Dutch National Pathology Databank (PALGA) and the Dutch Cancer Registry (IKNL) [12].

The model was updated to reflect insights into the natural history of cervical carcinogenesis [13]. Previously, the model contained a linear trajectory of disease progression (from HPV infection to CIN1, CIN2, CIN3 and cancer), while the current model describes progression from HPV infection through a persistent infection to a mixture of regressive and non-regressive lesions. The following new parameters were added to the updated model: the proportion of persistent infections developing into a non-regressive lesion, clearance rate of a regressive lesion and attributable fraction of CIN2 and CIN3 to regressive and non-regressive lesions. Furthermore, to arrive at a more parsimonious type-specific model, the natural history parameters for the 14 hrHPV types were clustered into four groups with similar rates of cancer progression: a) HPV-16, b) HPV-18, c) HPV-31, -33 and -45 (vaccine cross-protected types), and d) other hrHPV types.

The new parameters were calibrated, conditional on the values of the model parameters describing progression up to precancerous lesions, using a grid search method (Supplementary Appendix A.1.3, Supplementary Tables S.2-S.3, Supplementary Figures S.2-S.4). To do this, we used HPV type prevalence and CIN detection data from women with primary hrHPV testing in 2017-2018 with follow-up until 2019, i.e., shortly after the introduction of primary HPV-based screening in the Netherlands. Additionally, we used cancer incidence, averaged over the years 2015 and 2019 from the Dutch cancer registry [14], and HPV type attribution in cancer based on estimates from the literature [15].

### Dutch contemporary screening

The Dutch screening protocol was updated in July 2022, with hrHPV testing as the primary test and cytology and HPV16/18 genotyping as triage tests. Women with high-grade cytology (HSIL) or HPV-16/18 positive low-grade cytology (ASCUS/LSIL) are immediately referred for colposcopy. Other hrHPV-positive women are advised to repeat cytology after one year and are referred for colposcopy if they have abnormal cytology (ASCUS/LSIL/HSIL) at the repeat test. Non-referred women are re-invited for screening five or ten years after the primary test, depending on their HPV-test result and age.

Test characteristics are shown in Supplementary Table S.4 [16]. Based on the national screening monitor data in year 2022, we assumed that 15% of the women take part in the screening program through a self-sample test, while the other remaining 85% take the HPV-test at their primary care GPs office. For triage, we use a referral scheme based on HPV-16/18 genotyping, as currently implemented in the Dutch screening program, with the HPV-16/18 proportions among hrHPV positives estimated from POBASCAM data (Supplementary Appendix A.3) [11,17]. Demographic inputs, i.e. all-cause mortality and hysterectomy rates, were updated with the latest data (Supplementary Appendix A.4, Supplementary Table S.5-S.6) [18,19].

### HPV vaccination

In the base case we consider a vaccine efficacy of 95% for HPV-16 and HPV-18, and cross-protection efficacies of 75%, 50% and 80% for HPV-31, -33 and -45, respectively. We assume that vaccine efficacies have a lifetime duration. The cross-protective vaccine efficacies for non-vaccine types are based on empirical estimates in studies on the bivalent HPV vaccine [20–23]. Furthermore, the cohort-specific HPV vaccine coverage is based on reports from the Dutch vaccination registry (Supplementary Table S.7). Cohorts born between 1993-1995 were all vaccinated in 2009 as part of a catch-up campaign. All other birth cohorts (1996-2002) were vaccinated in the year they turned 13.

### Modelled screening strategies

The reference strategy is the current screening policy, with attendance rates based on the observed program attendance in 2022 (Supplementary Table S.8) [24]. To reiterate, according to current policy, all women are invited for HPV-based screening at fixed screening ages 30, 35, 40, 50 and 60 years, and women are invited at ages 45 and 55 years if they were HPV positive or did not attend in the previous round (five years earlier). Women are invited at age 65 years if they were HPV-positive at age 60 years and not referred to a gynecologist. The less intensive screening strategies considered here are consistent with the ages of invitation used in the current policy for ease of implementation.

We model eight alternative screening strategies, presented in Table 1. Strategies starting with N30 have no invitation at age 30, but start screening at age 35, with N30.1, N30.2 and N30.3 annotated in order of decreasing screening intensity from age 40 onward. Likewise, R35.1 and R35.2 are ordered by decreased screening intensity, with R35 denoting a risk-based round at age 35. This means that that women are invited only at age 35 if their HPV-test result was positive or they did not participate at age 30. Strategies N35 and N40 have no invitation at age 35 or at age 40, respectively, and have ten-year screening intervals thereafter. In the R strategy, fixed screening invitations are only sent at age 30; subsequent invitations from age 35 to 65 are all risk-based, meaning that women are only invited if their HPV-test result was positive or missing five years earlier.

**Table 1:** Screening policies: who is invited at what age?

| Strategy | Age <sup>a</sup> |  |  |  |  |  |  |  |
| --- | --- | --- | --- | --- | --- | --- | --- | --- |
|  | 30 years | 35 years | 40 years | 45 years | 50 years | 55 years | 60 years | 65 years |
| **Current** | All Women | All Women | All Women | HPV+ (previous round) | All Women | HPV+ (previous round) | All Women | HPV+ (previous round) |
| **N30.1** | No | All Women | All Women | HPV+ (previous round) | All Women | HPV+ (previous round) | All Women | HPV+ (previous round) |
| **N30.2** | No | All Women | HPV+ (previous round) | All Women | HPV+ (previous round) | All Women | HPV+ (previous round) | All Women |
| **N30.3** | No | All Women | No | All Women | No | All Women | No | All Women |
| **R35.1** | All Women | HPV+ (previous round) | All Women | HPV+ (previous round) | All Women | HPV+ (previous round) | All Women | HPV+ (previous round) |
| **R35.2** | All Women | HPV+ (previous round) | No | All Women | HPV+ (previous round) | No | All Women | No |
| **N35** | All Women | No | All Women | No | All Women | No | All Women | No |
| **R** | All Women | HPV+ (previous round) | HPV+ (previous round) | HPV+ (previous round) | HPV+ (previous round) | HPV+ (previous round) | HPV+ (previous round) | HPV+ (previous round) |
| **N40** | All Women | All Women | No | All Women | No | All Women | No | All Women |

<sup>a</sup> Women who were invited but did not attend the previous round five years earlier always get an invitation.

Additionally, we also consider strategies with less intensive screening applied only to vaccinated women, with unvaccinated women being screened according to the current policy. Thus, we considered a total of 16 alternatives to the current policy.

### Quality of life and costs

Utility values for the different health states and screening events are based on a Dutch quality of life study and previous cost-effectiveness analyses in the Netherlands [25,26]. Costs of screening and treatment are in euros (EUR), indexed to the year 2023 (Table 2). Costs of screening are based on the subsidy regulation to the cervical cancer screening program determined by the Dutch government [27]. The calculation of CIN follow-up and treatment costs were calculated from a population-based screening trial database [28], linked to the nationwide network and registry of histopathology (Supplementary Table S.9). Costs of cancer treatment are based on a previous publication and updated to 2023 prices [26].

**Table 2:** Model Inputs for health economic analyses.

|  | Total costs (€) | Disutility <sup>a</sup> | Duration (months) |
| --- | --- | --- | --- |
| **Screening tests <sup>b</sup>** |  |  |  |
| Primary hrHPV test | 44 | 0 | 0 |
| Primary hrHPV self-sampling kit | 26 | 0 | 0 |
| Reflex cytology after hrHPV-test | 30 | 0.03 | 1 |
| Repeat cytology after hrHPV self-sampling | 61 | 0.03 | 1 |
| Repeat cytology after 12 months | 61 | 0.03 | 12 |
| **Diagnosis and treatment <sup>c</sup>** |  |  |  |
| No CIN | 494 | 0.03 | 1 |
| CIN1 | 744 | 0.03 | 1 |
| CIN2 | 1571 | 0.03 | 1 |
| CIN3 | 1922 | 0.03 | 1 |
| FIGO1A | 6721 | 0.08 | 12 |
| FIGO1B | 15490 | 0.08 | 12 |
| FIGO2+ | 15610 | 0.14 | 12 |
| Cancer survivor | 0 | 0.03 | 120 |
| Palliative care | 35694 | 0.5 | 12 |

<sup>a</sup> Disutilities based on Dutch surveys [25,26]; <sup>b</sup> Screening test costs based on the subsidy regulation to the cervical cancer screening program determined by the Dutch government [27]; <sup>c</sup> CIN treatment cost calculation shown in Supplementary Appendix Table A.9. Cancer treatment costs based on a previous publication [26].

### Health impact and cost-effectiveness analysis

We model the first ten birth cohorts that are invited for HPV vaccination in the Netherlands, i.e., all women born between 1993 and 2002. Each woman is followed throughout her lifetime and we count outcomes from age 30, which is the start of the current screening program. We provide model-based projections of lifetime risk of cervical cancer, number needed to screen (NNS) and number needed to refer (NNR) to detect a CIN2/3 or cancer case (CIN2+). Furthermore, we present estimates of cervical cancer incidence under less intensive screening in unvaccinated women, and compare this to the cancer risk under the current screening policy in a cohort without vaccination.

We also provide projections of the quality adjusted life years (QALYs), the costs of screening and cancer treatment costs. QALYs are discounted at 1.5%, costs are discounted at 3% per year, and the cost-effectiveness threshold (CET) is set at EUR 20,000 per QALY in accordance with 2024 Dutch guidelines for preventive interventions [29]. To summarize the costs and health effects, we present the net monetary benefit (NMB), defined as:

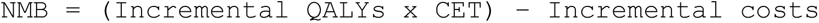

When using NMB, the current screening policy is the reference strategy (i.e., with NMB equal to zero). Strategies with an NMB larger than zero are cost-effective compared to the current screening policy and the policy with the highest NMB is considered the most cost-effective strategy. NMB is here presented in millions of euros per 100,000 women.

Furthermore, we also present the incremental cost-effectiveness ratio (ICER) defined as:

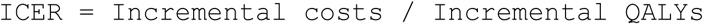

Here, each strategy is compared to next best strategy on the efficiency frontier, disregarding strategies that are dominated by other strategies (or combinations thereof) that lead to the same number of QALYs against lower costs. When using ICERs, a policy is considered cost-effective if the incremental cost per QALY is below the CET.

This study adheres to HPV-FRAME, a quality framework for mathematical modelling evaluations of HPV-related cancer control [30]. The checklist is reported in Supplementary Appendix Tables S.10-S.11.

### Sensitivity analyses

We repeat the analysis by (i) assuming 100% screening attendance and compliance with follow-up; (ii) using an international QALY set commonly used in modelling studies of cervical cancer screening [31,32]; (iii) presenting separate results for the last two birth cohorts in this analysis (birth years 2001-2002), which benefit the most from herd effects; (iv) presenting undiscounted results or using international discounting rates (3% for costs and health effects) as per WHO recommendation; and (v) considering less protection from vaccination by ignoring cross-protection against HPV-31, -33, or -45.

## RESULTS

### Effectiveness and efficiency of current screening strategy

Under the current screening strategy, the projected lifetime risk of cancer reduces from 633 per 100,000 women in cohorts without HPV vaccination, to 294 per 100,000 women in the first ten HPV-vaccinated birth cohorts. The NNS to detect CIN2+ increases from 64 to 130, and the NNR to detect a CIN2+ increases from 2 to 2.5. (Supplementary Figure S.5 and Tables S.12-S.13).

### Effectiveness and efficiency of less intensive screening strategies

Under the less intensive strategies considered, the number of cancers range from 304 to 569 cancers per 100,000 women (Figure 1). Thus, in all the strategies considered, the cancer incidence under less intensive screening would remain below the cancer incidence with current screening policy but without HPV vaccination. On the other hand, compared to the current policy without vaccination, the cancer risk in unvaccinated women would increase for all strategies with reduced screening intensity except for R35.1 (fixed screening rounds at ages 30, 40, 50 and 60, with risk-based invitations at ages 35, 45, 55 and 65) and R (screening starting at age 30 with risk-based invitations at five-year intervals afterwards) (Figure 1). In partly HPV-vaccinated cohorts, the efficiency of screening improves for less intensive strategies with NNS to detect CIN2+ ranging from 94 to 117, and the NNR to detect CIN2+ ranging from 1.8 to 2.4 (Supplementary Tables S.12 and S.13).

**Figure 1.**
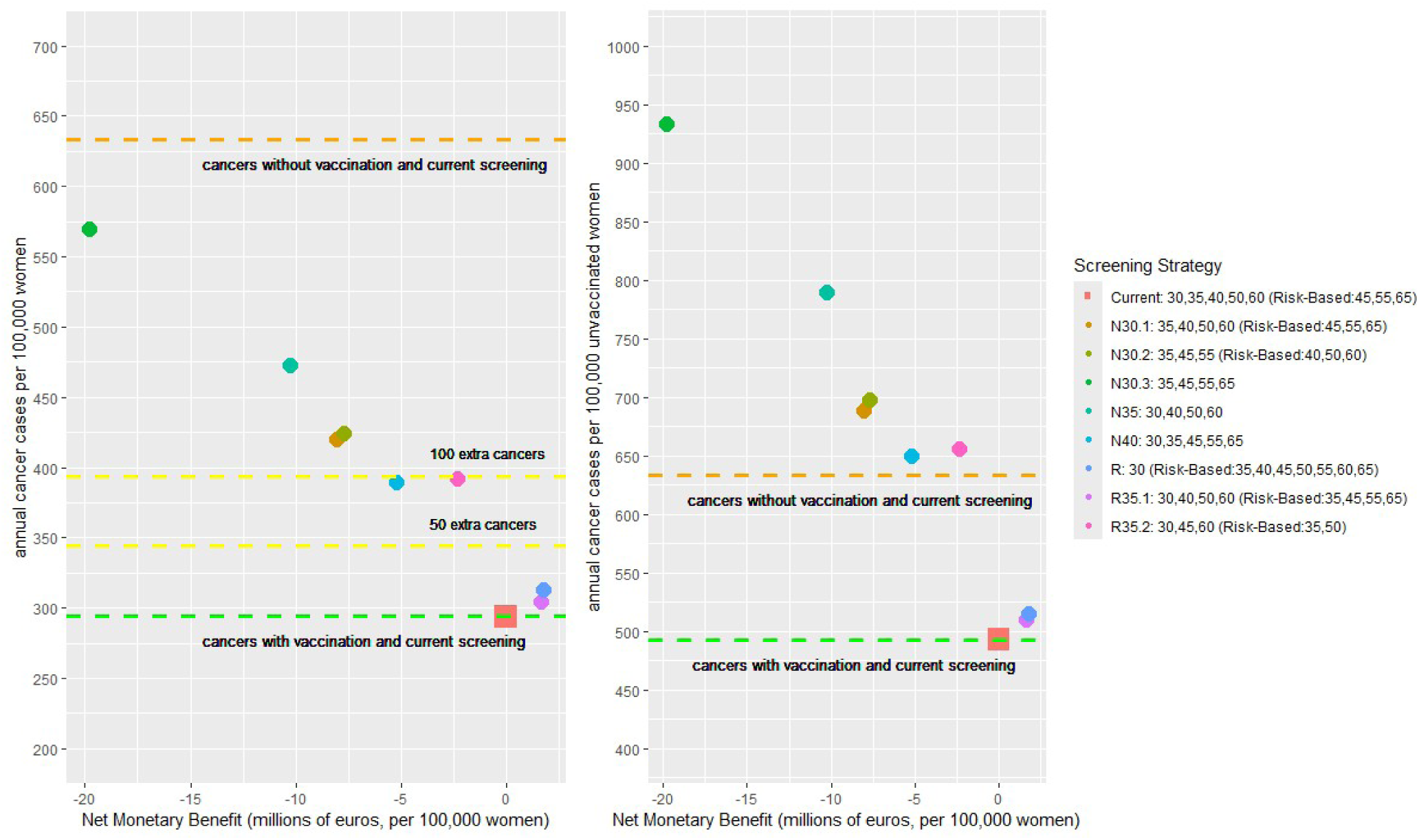
Net monetary benefit and cancer incidence relative to current policy. In the risk-based rounds, only women are invited who tested HPV-positive in the previous round or did not attend the previous round. Costs are discounted at 3% and QALYs are discounted at 1.5%.

### Net monetary benefit of less intensive screening strategies

The less intensive strategies result in cost savings between EUR 1.4 and 3.6 million per 100,000 women, with R35.2 (screening at ages 30, 45 and 60, with risk-stratified invitations at ages 35 and 50 years) being the least expensive strategy (Supplementary Table S.12). Strategies R35.1 and R result in costs savings of EUR 1.6 and 2.1 million, respectively, and would result in almost identical QALYs as compared to current screening policy of HPV-vaccinated cohorts, with 0 and 20 less QALYs per 100,000 women, respectively. On the other hand, all other strategies would result in much higher QALY reductions, ranging between 330 to 1160 less QALYs per 100,000 women as compared to current screening policy (Figure 1 and Supplementary Tables S.12-S.13).

In Figure 1, we show the cost-effectiveness of less intensive strategies using NMB. Strategies R35.1 and R have a positive NMB, meaning they are cost-effective compared to current screening, while all other strategies considered have a negative NMB. Strategy R35.1 yields EUR 1.6 million; and strategy R yields EUR 1.8 million per 100,000 women (Figure 1).

### Net monetary benefit of less intensive screening strategies stratified by HPV vaccination status

Strategies with stratification by HPV vaccination status result in monetary savings between EUR 0.9 and 2.3 million per 100,000 women, much lower than the savings without this stratification (Supplementary Table S.12). Figure 2 shows that strategy R again has the highest NMB, yielding EUR 1.3 million per 100,000 women. This value is lower than the maximum NMB where screening is reduced for all women. Hence, strategy R without stratification by HPV vaccination status is the most cost-effective of all 16 alternatives to the current policy considered here.

**Figure 2.**
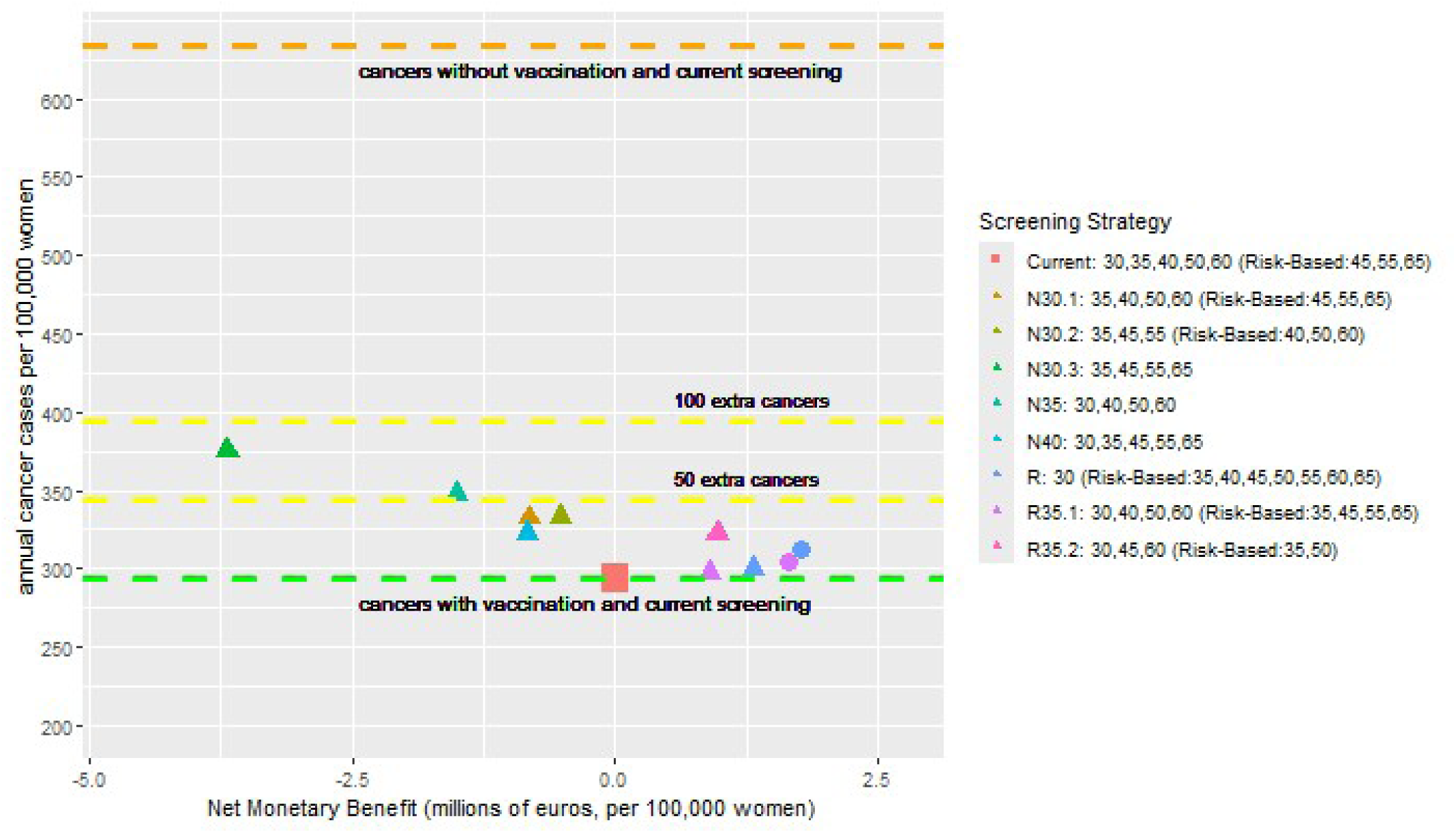
Net monetary benefit and cancer incidence relative to current policy, with screening intensity only reduced for vaccinated women. Triangles denote policies where screening is only changed for vaccinated women, while the circles denote the two best policies without stratification for HPV vaccination status. Risk-based rounds only apply to women who did not test HPV-negative in the previous round. Costs are discounted at 3% and QALYs are discounted at 1.5%.

### ICER of less intensive screening strategies

Figure 3 shows the costs and QALYs of all 16 screening strategies, with screening intensity reduced for either all or only HPV-vaccinated women. R35.2 (all women) is the least expensive strategy, and three strategies are further located on the efficiency frontier: R (all women), R35.1 (all women) and R (vaccinated women only). The corresponding ICERs of these strategies are EUR 5,543, 25,965 and 90,809 per additional QALY gained, respectively. At a CET of EUR 20,000 per QALY, the latter strategy cannot be considered cost-effective, whereas the choice between R and R35.1 (all women, in either case) hinges on the strictness of the CET.

**Figure 3:**
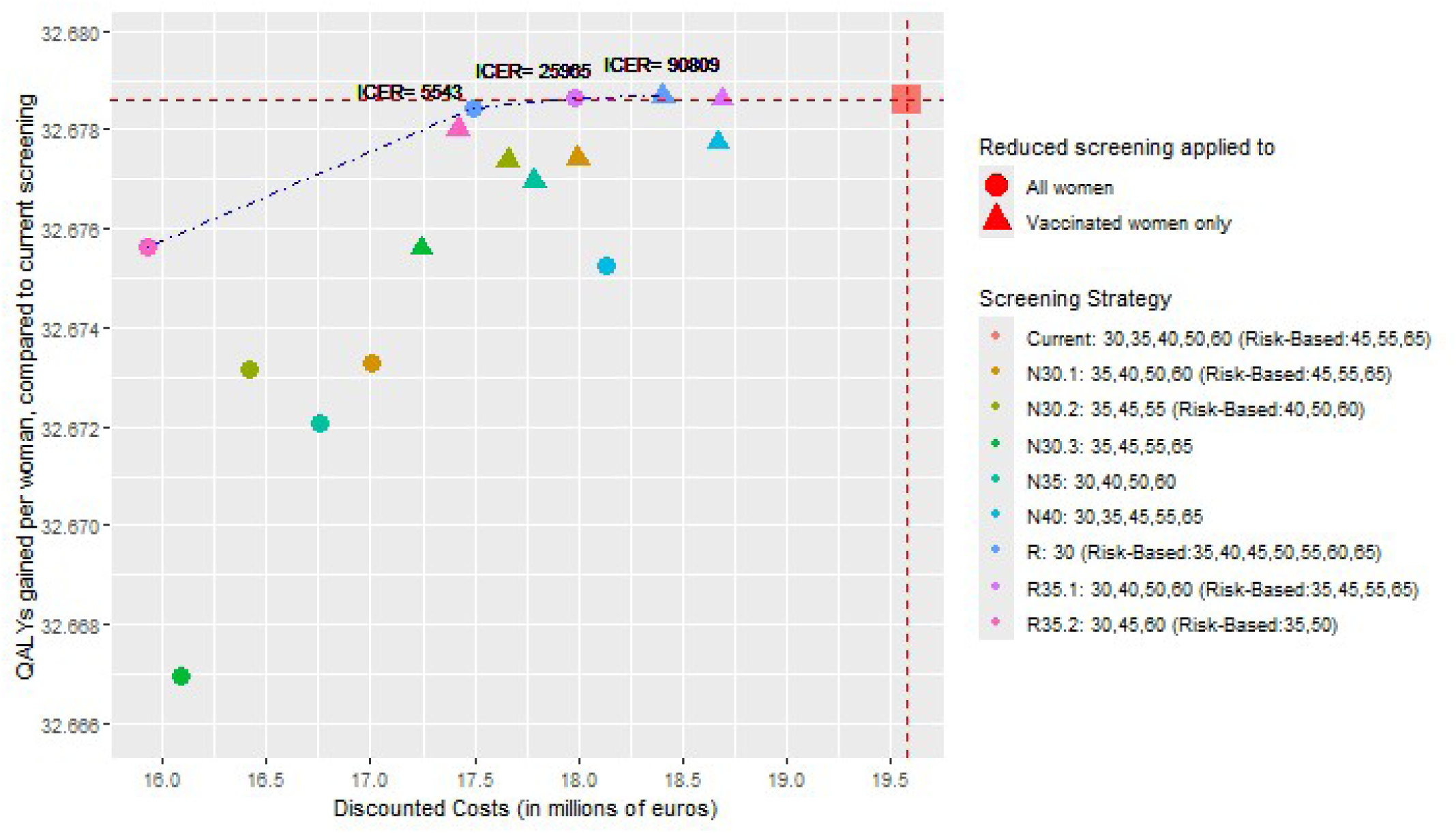
Costs and health gains of reduced screening in partly HPV-vaccinated cohorts. Triangles denote policies where screening is only changed for vaccinated women, while circles denote policies without stratification for HPV vaccination status. Risk-based rounds only apply to women who did not test HPV-negative in the previous round. Costs are discounted at 3% and QALYs are discounted at 1.5%.

### Sensitivity analyses

Figure 4 shows the sensitivity analyses for NMB of the three screening policies on the efficiency frontier in Figure 3.

**Figure 4:**
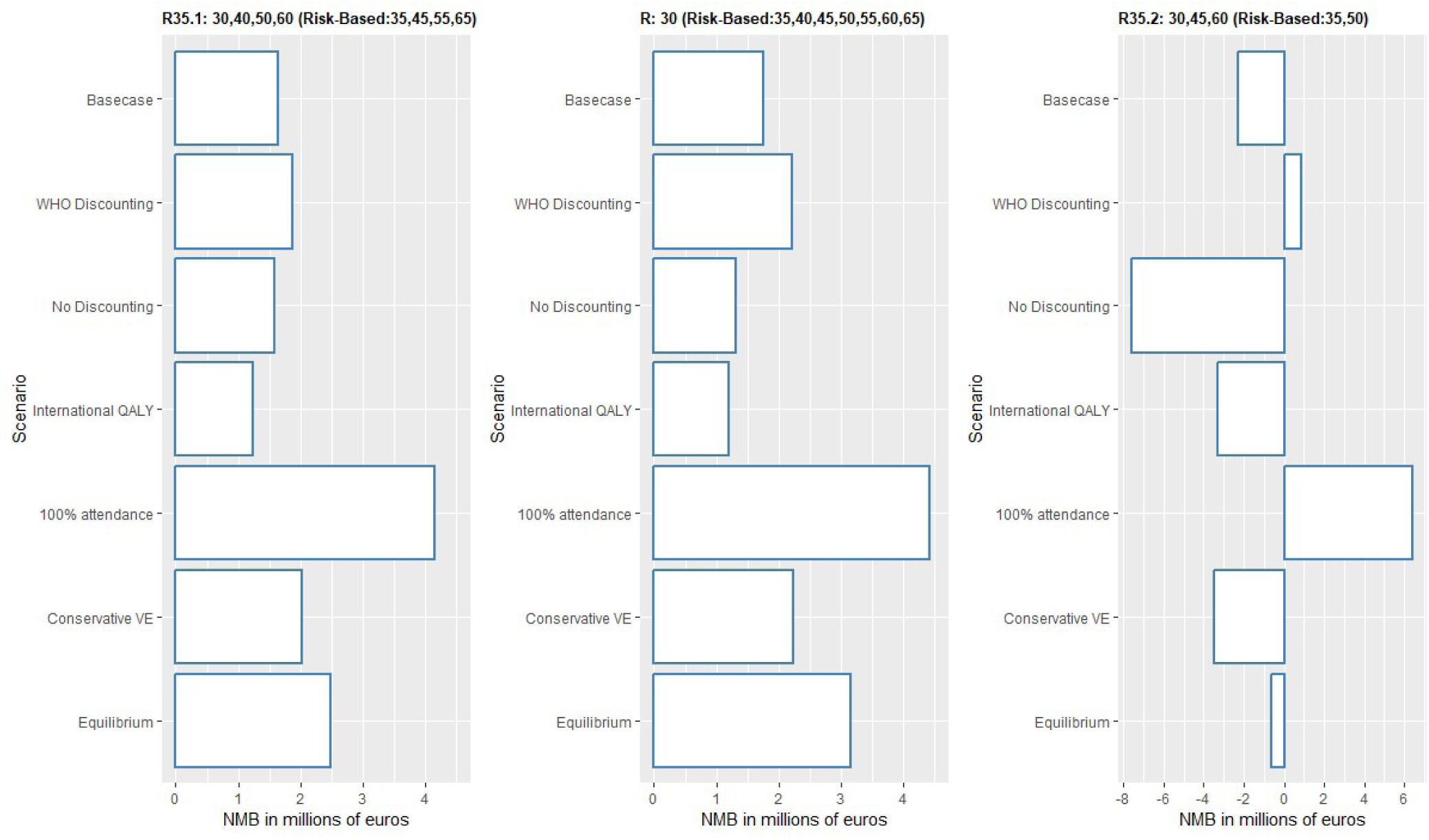
Sensitivity analyses. NMB denotes net monetary benefit per 100,000 women. WHO discounting consists of 3% discounting rates for costs and health effects. International QALY analysis is based on a set of utility weights previously used in cervical cancer modelling studies [31,32]. 100% attendance includes 100% compliance with all follow-up procedures. Conservative VE (vaccine efficacy) denotes no cross-protection from HPV-16/18 vaccination. Equilibrium is based on an analysis of the 2001 and 2002 birth cohorts, for whom the indirect effects of vaccination approached an equilibrium state.

If attendance and compliance with follow-up are set at 100%, more policies of reduced screening intensity would result in QALYs gained compared to current screening (Supplemental Figures S.7 and S.8). The strategy with the highest NMB is then R35.2, yielding EUR 6.4 million per 100,000 women. However, this strategy would result in a higher cancer risk in unvaccinated women than in the pre-vaccination situation (Supplementary Figures S.8). Conversely, the cancer risk in unvaccinated women remains reduced both under R35.1 and R, with NMB equal to EUR 4.1 million 4.4 million, respectively, per 100,000 women (Figure 4 and Supplementary Figures S.6 and S.7).

Other sensitivity analyses (including separate analysis of women born in 2001-2002) did not significantly change the results. However, in some scenarios (no discounting, alternative set of QALY weights) strategy R35.1 had the highest NMB instead of strategy R (Figure 4 and Supplementary Figures S.6-S.11). This is due to the relatively small difference in QALYs between strategies R35.1 and R, by which a slightly different weighting of quality loss due to screening procedures reverses their order in cost-effectiveness analysis.

## DISCUSSION

In this modelling study we investigated whether cervical cancer screening intensity can be reduced for the first ten birth cohorts that have been offered HPV vaccination in the Netherlands. Our study indicates that the starting age of screening can be maintained at 30 years of age, both for unvaccinated and vaccinated women, but screening above age 30 could become less intensive. A screening invitation at age 35 is only needed for women who do not test HPV-negative five years earlier, similar to the risk-based invitations at age 45, 55 and 65 in the current Dutch screening program. Furthermore, risk-based invitations could be expanded to all rounds, thereby abandoning the principle of fixed screening rounds where invitations are sent to all women.

Importantly, these risk-stratified strategies do not increase the cervical cancer risk for unvaccinated women in a setting with moderate vaccination coverage, as compared to a situation without HPV vaccination. By contrast, strategies starting age of 35 instead of 30 years, or strategies where screening takes place in standard ten-year intervals, would lead to an increased risk of cervical cancer in unvaccinated women. This is because vaccination coverage ranged from only 49% to 62% in the first ten birth cohorts in the Netherlands. Our findings are robust to assumptions regarding utility weights, discount rates, HPV vaccine cross-protection and the degree of indirect protection, as shown by separate analysis of later birth cohorts. In a setting with perfect screening attendance, an even less intensive strategy might be preferred, however such a policy would also lead to an increased cancer risk for unvaccinated women.

We also found that policies with stratification for HPV vaccination status are less cost-effective than policies without such stratification. Although stratification ensures that unvaccinated women are not at increased risk of cancer, our results show that they may also benefit from less intensive screening. Furthermore, implementing a screening policy stratified for HPV vaccination status is not straightforward in the Netherlands, since the Dutch screening and vaccination registries are not coupled and linkage of registries is logistically complex and subject to privacy rules.

Additionally, such policies could complicate communication to the public; for instance, some vaccinated women might interpret the reduction in screening intensity as a downgrade in their access to health services. Instead, re-inviting women who test positive for HPV after five instead of ten years is more easily communicated, namely as an extra health service for those who are at increased risk.

Multiple mathematical modelling studies previously examined the benefits and harms of cervical cancer screening, and we found six studies on cervical cancer screening of vaccinated cohorts [5,6,31,33–35]. All studies agreed that with the introduction of HPV vaccination, de-intensification of screening would be necessary to ensure cost-effectiveness. However, most studies started from a setting with higher screening intensity and higher vaccination rates than currently applies to the Netherlands, and none of these studies considered risk-stratified screening, with optional screening rounds based on HPV-test outcome or attendance in previous rounds. Only one study from Norway considered stratification on the basis of HPV vaccination status [6]. This study found, similar to us, that reduced screening of HPV-vaccinated women would be more efficient than continuation of the current screening policy, but no comparison with risk-based strategies for all women, irrespective HPV vaccination status, was performed.

Our study has several strengths. We decided to only consider a limited set of screening policies after discussion with relevant stakeholders. The strategies evaluated here retain the same ages for invitation as in the current program, which facilitates implementation. The relatively low vaccination coverage combined with reduced screening intensity could generate a substantial inequality in cancer risk between vaccinated and unvaccinated women. We have taken these equity concerns into account by also presenting the model estimates of cancer risk in unvaccinated women, and adopting the constraint that these should not be higher than under the current screening policy without vaccination [36].

In this study we assessed cost-effectiveness both by ICER and by NMB. Given the small difference in QALYs, NMB results in more stable estimates compared to ICER. For instance, the ICER for extending risk-based invitations to every screening round is substantially lower than extending risk-based invitations to age 35 only, however, when we look at NMB, we verify that these two strategies are relatively close to each other in terms of cost-effectiveness. Furthermore, NMB facilitates the cost-effectiveness calculation in a setting where different constraints are possible (e.g., cancer risk in unvaccinated women or cancer risk increase in all women) when deciding between screening strategies [37].

Our study also has some limitations. First, the degree of indirect protection provided by HPV vaccination was based on an HPV transmission model, and not yet on empirical estimates. Second, our model assumes that HPV prevalence and cervical cancer incidence would remain stable over time in the absence of vaccination. This assumption is convenient from a computational perspective, and a necessary simplification because it is difficult to extrapolate historic trends in HPV prevalence and cancer incidence into the future. Third, we assume no association between vaccination status and screening attendance, which could lead to an underestimation of cancer incidence with less intensive screening, if unvaccinated women have a lower inclination to participate in screening.

While the recommendations provided here are derived from the Dutch setting, the de-intensification of screening considered in this modelling study contains valuable lessons for other countries where HPV-vaccinated women are entering the screening program. Some European countries have a vaccination coverage that is similar (Germany) or lower (France) than that in the Netherlands [8], and for these it would seem pertinent to consider the cancer risk for unvaccinated women as a way to constrain the set of possible reduced screening policies.

Additionally, risk-stratified screening, based on previous screen attendance and prior HPV-test outcomes, could be a promising approach for other European countries as well. A possible avenue for further risk stratification could be to use multiple screening rounds instead of only the previous round to inform a woman’s risk of cervical cancer [38].

In conclusion, with the entrance of HPV-vaccinated birth cohorts, cervical cancer screening in the Netherlands could become less intensive by inviting women after ten years following a negative HPV-test or after five years otherwise. Alternatively, fixed screening ages with ten-year intervals could be kept (at ages 30, 40, 50 and 60) with selective screening invitations for the in-between rounds. Further de-intensification of the screening program is premature, as this could lead to an increased cancer risk for the unvaccinated women.

## Data Availability

All data produced in the present study are available upon reasonable request to the authors.

## Author contributions

**Tiago M. de Carvalho:** Conceptualization; methodology; data curation; simulation; formal analysis; writing – original draft. **Johannes Berkhof:** Funding acquisition; conceptualization; methodology; writing – review and editing. **Johannes A. Bogaards:** Funding acquisition; conceptualization; methodology; writing – review and editing.

