## Supplementary material for "Cost-effectiveness of risk-stratified screening for cervical cancer in cohorts vaccinated against human papillomavirus with moderate vaccination coverage": Supplemantary Material

**List of Figures**

Figure S.1: Natural history from HPV infection to cervical cancer.

Figure S.2: Model fit for HPV prevalence of HPV-16, HPV-18 and other types.

Figure S.3: Model fit for CIN detection.

Figure S.4: Model fit for cervical cancer incidence.

Figure S.5: Cancer incidence with vaccination by birth cohort and current screening policy.

Figure S.6: Costs and health gains of reduced screening in partly HPV-vaccinated cohorts under 100% screen attendance.

Figure S.7: Net monetary benefit and cancer incidence relative to current policy without vaccination under 100% screen attendance.

Figure S.8: Costs and health gains of reduced screening in partly HPV-vaccinated cohorts under alternative QALY weights.

Figure S.9: Net monetary benefit and cancer incidence relative to current policy without vaccination under alternative QALY weights.

Figure S.10: Costs and health gains of reduced screening in partly HPV-vaccinated cohorts under conservative vaccine efficacy.

Figure S.11: Net monetary benefit and cancer incidence relative to current policy without vaccination under conservative vaccine efficacy.

**List of Tables**

Table S.1: List of model parameters.

Table S.2: Grid search of natural history parameters.

Table S.3 Grid search of CIN attribution parameters.

Table S.4: Test characteristics of the hrHPV test and cytology by health state.

Table S.5: Hysterectomy rates by age group.

Table S.6: Survival from diagnosis by cervical cancer stage.

Table S.7: Vaccination coverage in the Netherlands by birth cohort.

Table S.8: Screening attendance in the Netherlands by five-year age group.

Table S.9: Calculation of CIN follow-up and treatment costs.

Table S.10: HPV-FRAME Checklist, Core Reporting Standard.

Table S.11: HPV-FRAME Checklist, Reporting standard for HPV vaccination in adolescent individuals.

Table S.12: Health and economic outcomes of cervical cancer screening in partly HPV-vaccinated cohorts.

Table S.13: Additional undiscounted model outcomes for current screening and alternative screening policies in partly HPV-vaccinated cohorts.

**Supplementary Appendix A: Technical Appendix**

### A.1 HPV transmission model and cancer progression model

#### A.1.1 HPV transmission model

The HPV transmission model is a deterministic compartmental model for heterosexual partnership formation, using a SIRS (Susceptible, Infected, Recovered/Immune, Susceptible) structure and is extensively described in Bogaards et al. 2010.[1] The model describes 14 oncogenic types, namely HPV genotypes 16, 18, 31, 33, 35, 39, 45, 51, 52, 56, 58, 59, 66 and 68. The model considers three levels of gender-specific rates of partner change corresponding to low, intermediate and high levels of sexual activity. The model distinguishes between primary and persistent stages of HPV infection in women, and contains a description of natural history up to development of precancerous cervical lesions in women. HPV type-specific model parameters were estimated from POBASCAM data, a prospective randomized controlled trial of HPV testing for cervical screening, using a Bayesian framework with an MCMC algorithm.[1,2] For this study we reduced the number of parameters from 14-type specific sets to four groups of epidemiologically similar types by weighted pooling, namely: a) HPV-16, b) HPV-18, c) HPV-31, -33, -45 (types for which cross-protection from HPV-16/18 vaccination is assumed) and d) all other oncogenic types (types for which no cross-protection is assumed). Weights for pooling were defined based on oncogenicity according to de Sanjose et al. 2010.[3] The posterior means of the parameter estimates were used as input for the cancer progression model (Table S.1), which contains a further description of cervical carcinogenesis, from infection to screen-detectable CIN lesions to invasive cervical cancer.

#### A.1.2 Cancer progression model

The cancer progression model is an individual-based discrete-time microsimulation model that simulates birth cohorts of women from age 10 to death, in time steps of six months. The part of the model that describes the natural history until invasive cancer consists of 14 parallel Markov chains, corresponding to the different oncogenic HPV types. To project the effects of various screening strategies in women born between 1993 until 2002, we constructed six-month age-birth cohorts with corresponding probabilities of type-specific HPV infection predicted by the HPV transmission model. To this end, we transformed the force of infection from a continuous time hazard to a six-month cumulative probability in discrete time for use in the cancer progression model, conditional on the level of sexual activity.

The structure of the cancer progression model with regard to the natural history of type-specific HPV infection conforms to that of the HPV transmission model, up to development of precancerous cervical lesions for each of the 14 high-risk HPV types. The cancer progression model also contains a type-specific immune state that follows upon clearance of infection, with duration $\mu_i$ (where $i$ denotes the HPV type), estimated as part of the transmission model.[1] The duration of infection follows a type-specific distribution with six (discrete-time) transition parameters $\gamma_i, \eta_i, \delta_{i,1}, \delta_{i,2}, \nu_{i,1}, \nu_{i,2}$. Some of these parameters were estimated in Bogaards et al. 2010 [1] and also used in Bogaards et al. 2011 [4] and Berkhof et al. 2013 [5] namely: $i$) $\eta_i$ denoting the type-specific progression rates from incident to persistent HPV infection; $ii$) $\gamma_i$ and $\delta_{i,1}$

denoting the type-specific clearance rates for incident and persistent HPV infections. Note that persistent HPV infections may still clear, but do so at a much smaller rate than incident HPV infections (i.e. $\gamma_i > \delta_{i,1}$ for all types *i*). These parameters are shared between the HPV transmission model and the cancer progression model (after transformation from rates in continuous time to state transition probabilities in discrete time).

Compared to previous studies we modified the structure of the cancer progression model after HPV persistence. The previous model contained a linear trajectory from cervical intraepithelial neoplasia (CIN) grade 1, 2 and 3 to invasive cancer. The modified structure assumes that the infection first becomes persistent after which the infection can progress to a regressive or non-regressive lesion. On a histo-morphological level, a new HPV infection corresponds with CIN0, a persistent infection with CIN0/1, regressive lesions with CIN1/2/3, and non-regressive lesions with CIN2/3. This new structure better reflects epigenetic insights into cervical carcinogenesis.[6] Furthermore, it facilitates re-calibration to new data from primary HPV-testing in the Dutch cervical cancer screening program. By separating diagnostic categories from the underlying natural history of cancer progression, we also increased the flexibility of the model to match changes in diagnostic criteria and performance.

To elaborate on the modified model structure, a persistent HPV infection may either clear (with six-month transition probability $\delta_{i,1}$), or transition to a precancerous lesion, with six-month transition probability $v_i$,

$$v_i = \frac{\mathrm{N}_i}{\mathrm{N}_i + \Delta_{1i}} * (1 - \exp(-(\mathrm{N}_i + \Delta_{1i}) * 0.5)) .$$

The hazards $\mathrm{N}_i$ and $\Delta_{1i}$ underlying these probabilities were estimated as part of the HPV transmission model in Bogaards et al. 2010 [1] and remain unchanged in the model presented here. For the purpose of this study, the transition to precancerous lesions was further distinguished into progression to either a regressive lesion ($v_{i,1}$ ) or a non-regressive lesion ($v_{i,2}$). Transition parameter $v_{i,1}$ is defined as $(1 - p_i)v_i$ and conversely $v_{i,2}$ is defined as $p_i v_i$. The parameter $p_i$ thus indicates the type-specific proportion of persistent infections that develop into non-regressive lesions vs. regressive lesions.

Finally, regressive lesions may clear with six-month type-specific probability $\delta_{i,2}$, which is defined as,

$$\delta_{i,2} = 1 - \exp(\Delta_{2i} * 0.5 ).$$

In this study, $p_i$ and $\delta_{i,2}$ were estimated for the four grouped sets of HPV types (HPV-16, HPV-18, HPV-31/33/45, other high-risk types) during re-calibration of our hybrid model to outcomes of primary HPV-based screening program in the Netherlands (see section A.1.3 below). Finally, conditional on these estimated parameter values, two new parameters for the attribution of CIN1, 2 and 3 lesions to regressive and non-regressive lesions were fitted, assuming that CIN1 lesions can only be regressive and that only 10% of the non-CIN1 regressive lesions are CIN3.[7] The newly introduced parameters *prop.cin1* and *prop.cin3* denote the proportion of CIN1 in regressive lesions and the proportion of CIN3 in non-regressive lesions, respectively.

The duration between the onset of a non-regressive lesion and invasive cervical cancer (denoted as $G(k_{i,} \theta_i)$ in Figure A.1) was modelled as a gamma distribution, with $k_i$ denoting the shape parameter and $\theta_i$ the scale parameter (distinct for HPV types 16 and 18 versus other high-risk types). These parameters were previously estimated by linking the age distributions of CIN2/3 to cervical cancer based on data from the Dutch National Pathology Databank (PALGA) and the Dutch cancer registry.[8] Progression between invasive cancer states FIGO1a, FIGO1b and FIGO2+ was based on national registry and screening data (Figure S.1, Table S.1).

**Figure S.1: Natural history from HPV infection to cervical cancer.**

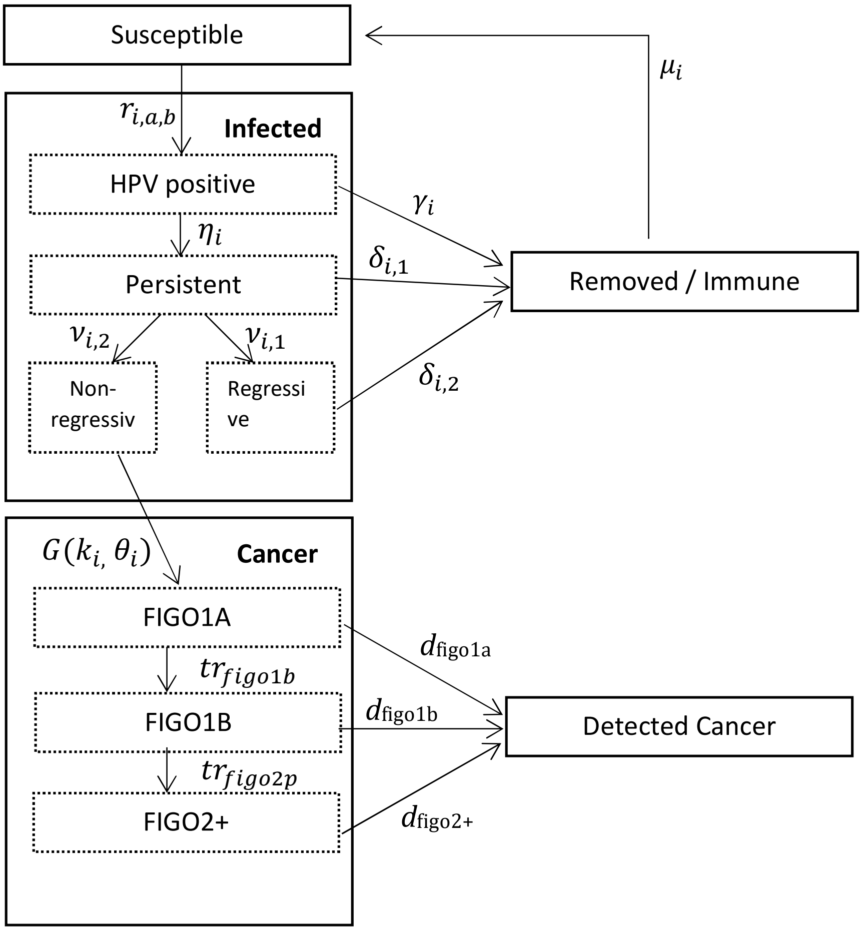

**Table S.1: List of model parameters.**

| Notation | Description | Values by HPV type: 16; 18; CP; NCP [a] | Reference |
|---|---|---|---|
| ***Natural history of infection*** [b] | | | |
| $r_{i,a,b,l}$ | Forces of infection for HPV-16, HPV-18, cross-protected types and non-cross-protected types. | Range of values per HPV type (*i*) , age group (*a*) ,birth cohort (*b*) and sexual activity level (*l*). | Computed using HPV transmission model. |
| $\gamma_{16}, \gamma_{18}, \gamma_{cp}, , \gamma_{ncp}$ | Clearance rate from incident HPV infection. | 0.29; 0.33; 0.37; 0.44 | Bogaards et al. 2010 [1] |
| $\eta_{16}, \eta_{18}, \eta_{cp}, \eta_{ncp}$ | Progression rate from incident to persistent HPV infection. | 0.24; 0.19; 0.16; 0.09 | Bogaards et al. 2010 [1] |
| $\delta_{16,1}, \delta_{18,1}, \delta_{cp,1}, \delta_{ncp,1}$ | Clearance rate from persistent infection. | 0.06; 0.17; 0.16; 0.24 | Bogaards et al. 2010 [1] |
| $\delta_{16,2}, \delta_{18,2}, \delta_{cp,2}, \delta_{ncp,2}$ | Clearance rate from regressive lesions. | 0.11; 0.12; 0.07; 0.06 | Section A.1.3 |
| $\nu_{16,1}, \nu_{18,1}, \nu_{cp,1}\ \nu_{ncp,1}$ | Progression rate from persistent infection to regressive lesions. | 0.13; 0.09; 0.13; 0.07 | Section A.1.3 |
| $\nu_{16,2}, \nu_{18,2}, \nu_{cp,2}\ \nu_{ncp,2}$ | Progression rate from persistent infection to non-regressive lesions. | 0.06; 0.09; 0.01; 0.01 | Section A.1.3 |
| $\mu_{16}, \mu_{18}, \mu_{cp}, \mu_{ncp}$ | Rate of waning natural immunity. | 0.02; 0.01; 0.02; 0.02 | Bogaards et al. 2010 [1] |
| ***Duration to invasive cervical cancer*** | | | |
| $k_{16}, k_{18}, k_{cp}, k_{ncp}$ | Shape parameter of gamma distribution. | 9.67; 9.67; 2.49; 2.49 | Vink et al. 2013 [9] |
| $\theta_{16}, \theta_{18}, \theta_{cp}, \theta_{ncp}$ | Scale parameter of gamma distribution. | 3.33; 3.33; 9.14; 9.14 | Vink et al. 2013 [9] |
| ***Cervical cancer progression & detection*** | | | |
| $d_{figo1a}, d_{figo1b}, d_{figo2p}$ | Detection rates for FIGO1a/FIGO1b/FIGO2+ health states in absence of screening. | 0.025; 0.05; 0.3 | Coupe et al. 2012 [10] |
| $tr_{figo1b}, tr_{figo2+}$ | Transition rate from FIGO1a to FIGO1b and from FIGO1b to FIGO2+ | 0.125; 0.1 | Coupe et al. 2012 [10] |

[a] *Parameters for natural history and duration to cancer are separately given for HPV-16; HPV-18; cross-protected (CP) types, i.e. HPV-31, -33 and -45; and types not covered by cross-protection (NCP), namely HPV-35, -39, -51, -52, -56, -58, -59, -66 and -68.*

[b] *These parameters are shared between the transmission model and the cancer progression model. All rates are cumulative probabilities per 6-month period, unless otherwise indicated.*

#### A.1.3 Model calibration

**Overview**

For this study we first calibrated the two newly defined parameters in the natural history, $\delta_{i,2}$ and $p_i$ . Conditional on these parameter values we calibrated *prop.cin1* and *prop.cin3* to screening data. For this we used data from the first round of the Dutch screening program after the introduction of primary HPV testing, spanning a period of screening invitations between 2017 and 2018, with follow-up until 2019. From this dataset, we distilled 49 calibration targets, including HPV prevalence between ages 30 and 60, subdivided by HPV-16, HPV-18 and other high-risk HPV types, CIN detection between ages 30 and 60, stratified by CIN1, CIN2, and CIN3, and cancer incidence between ages 20 and 65.[8] Additionally, we used published estimates from de Sanjose et al. 2010 [3] to match HPV type attribution in cancer, grouped by a) HPV-16, b) HPV-18, c) HPV-31/33/45, d) other high-risk types.

**Method description**

We used a grid search method to find parameter sets consistent with the observed data. The algorithm is described as follows. Before starting the grid search, we set a range for each model parameter, based on expert opinion and previous versions of the model.[1] For example, we know a priori, based on type attribution data that the proportion non-regressive lesions of HPV-16 infections should be substantially higher than for all other types.[3] To identify whether the model prediction is within the limits for the corresponding calibration target, we define an error tolerance limit ($\epsilon$). We set *S* defined as the number simulation model runs to be performed. We also define a decision rule (*D*) to determine whether a sampled parameter is accepted as consistent with the calibration targets, for instance, a) the model prediction has an error smaller than $\epsilon$ for 80% of the calibration targets; and b) within one category no more than a single calibration target has an absolute error 2 times $\epsilon$, to exclude very poor fits.

Then, for each calibration round, we repeat the following steps,

1. Generate *S* model parameter values using Latin hypercube sampling (R package: *lhs*)
2. Run the cancer progression model *S* times using the parameters in Step 1.
3. In each $s$-th run compute the absolute value of the prediction error for every calibration target and compare it with $\epsilon$ and determine whether the $s$-th sampled parameter is accepted based on *D*.

These calibration rounds can be repeated several times until a region of the parameter space is found where enough parameters satisfy a target (e.g. 85% of the calibration targets are satisfied). In a final run, with a relatively large *S'* number of runs compared to *S* we determine the "optimum" parameter set. In case there are several parameter combinations satisfying the same number of calibration targets, we choose the model parameter combination that minimizes the absolute error in cancer incidence.

Once the optimal $\delta_{i,2}$ and $p_i$ are found, the CIN attribution parameters are optimized. Since these parameters only affect the model prediction for CIN detection, only observed CIN data was used for the latter optimization. The optimal *prop.cin1* is found as the one with the lowest mean absolute error in predicted CIN1, with the proportion of CIN2 diagnoses given by (1- *prop.cin1),* and similarly for *prop.cin3*, conditional on the optimum *prop.cin1,* by finding the *prop.cin3* which results in lowest mean absolute error in predicted CIN3.

**Model calibration results**

The initial grid per model parameter is shown in Table S.2-S.3. We chose $S$=200 for the first waves. We considered that at least 85% of the calibration targets would need to be satisfied as a stopping rule for the grid search. We achieved this after two waves. Then we ran the model $S'$=1000, for the last run. We set tolerance error $\epsilon$ as 20% times the calibration target, with an exception for all targets corresponding to proportions where the tolerance error was defined as $max$(20%, 0.005). This means that for instance, for the rate of CIN detection we accept a maximum error of either 20% of the observed CIN prevalence or an absolute error of 0.005 in relation to the observed CIN prevalence. The results of the grid search are also shown in Tables S.2-S.3. The model fit is shown Figures S.2-S.4.

**Table S.2 Grid search of natural history parameters.**

| **Model Parameter** | $p_{16}$ | $p_{18}$ | $p_{cp}$ | $p_{ncp}$ | $\Delta_{2,16}$ | $\Delta_{2,18}$ | $\Delta_{2,cp}$ | $\Delta_{2,ncp}$ |
|---|---|---|---|---|---|---|---|---|
| **Initial Range** | | | | | | | | |
| Lower bound | 0.15 | 0.01 | 0.01 | 0.01 | 0.15 | 0.15 | 0.1 | 0.1 |
| Upper Bound | 0.40 | 0.15 | 0.15 | 0.15 | 0.45 | 0.45 | 0.4 | 0.4 |
| **Optimal Parameters** | | | | | | | | |
| Optimum | 0.28 | 0.10 | 0.07 | 0.06 | 0.27 | 0.25 | 0.15 | 0.13 |

**Table S.3 Grid search of CIN attribution parameters.**

| **Model Parameter** | $prop.cin1$ | $prop.cin3$ (regressive) | $prop.cin3$ (non-regressive) |
|---|---|---|---|
| **Initial Range** | | | |
| Upper bound | 0.3 | Fixed | 0.3 |
| Lower Bound | 0.9 | Fixed | 0.9 |
| **Optimal Parameters** | | | |
| Optimum | 0.59 | 0.1 x *(1-$prop.cin1$)* | 0.78 |

**Figure S.2 Model fit for HPV prevalence of HPV-16, HPV-18 and other types**

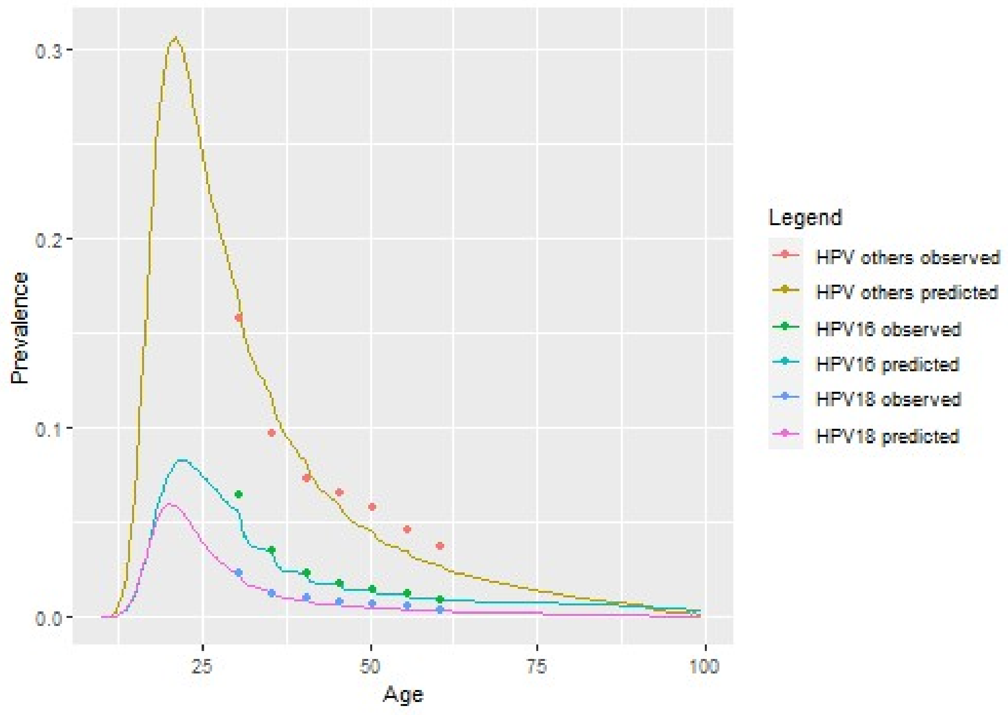

**Figure S.3 Model fit for CIN detection**

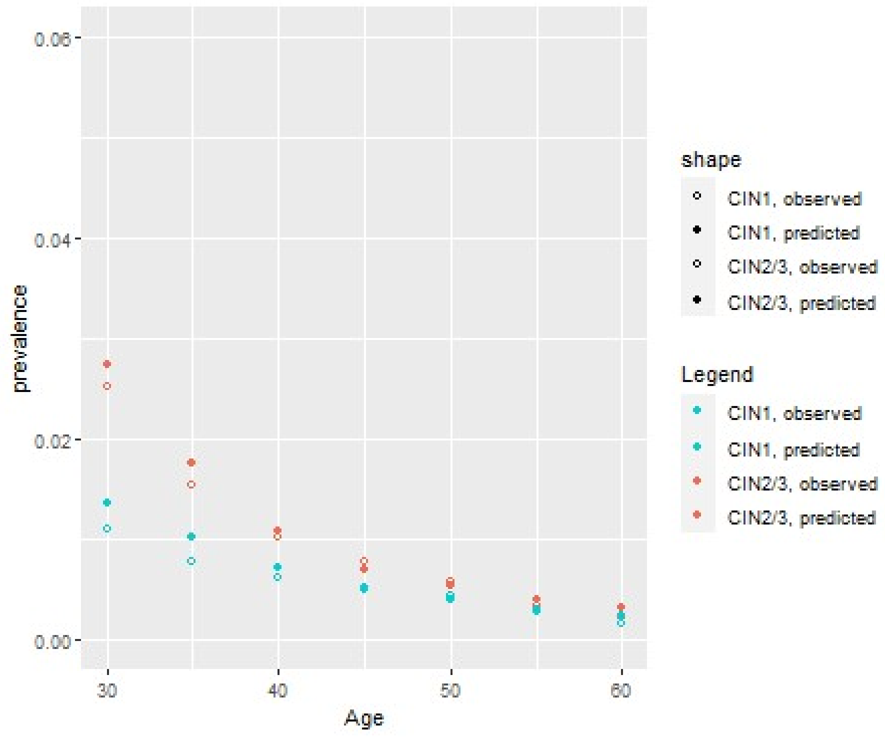

**Figure S.4 Model fit for cervical cancer incidence**

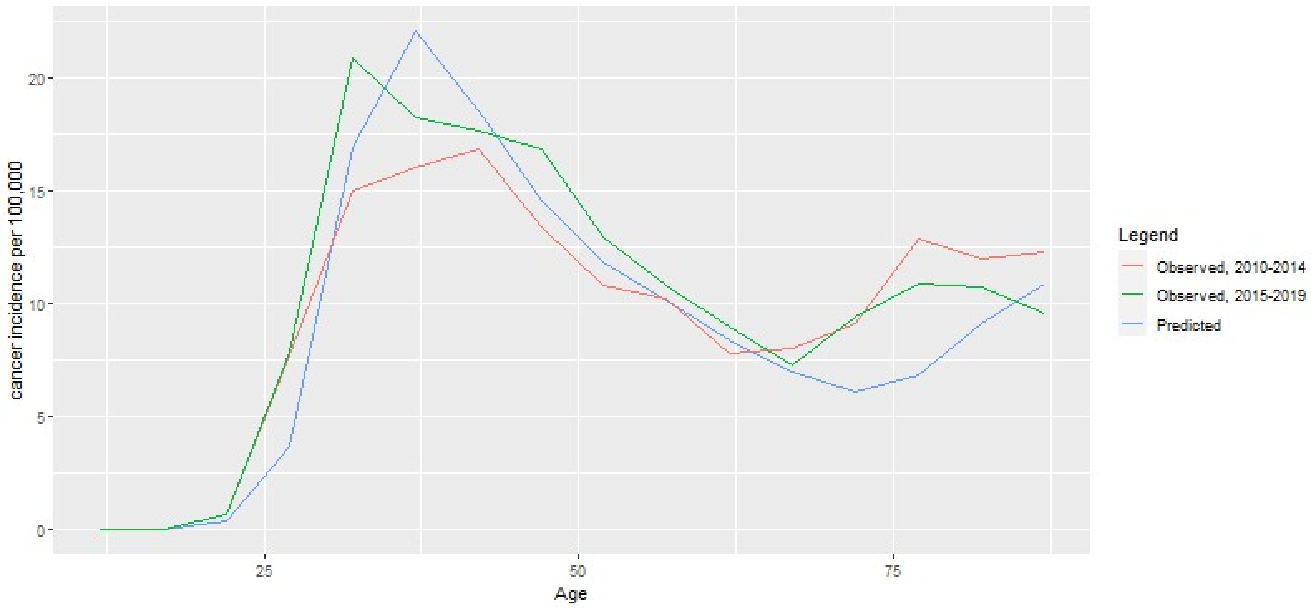

**Table S.4: Test characteristics of the hrHPV test and cytology by health state.**

| Test Result and Health State | Probability of Test Result |
|---|---|
| Positive hrHPV-test, in absence of hrHPV infection | 0% |
| Positive hrHPV-test, if hrHPV infection is present | |
| No CIN/ CIN1 | 70% |
| CIN2+ | 94% |
| Cytology ≥ Pap2,[a] if no hrHPV infection is present | 1.3% |
| Cytology ≥ Pap2, if hrHPV infection is present | |
| - No CIN | 5% |
| - CIN1 | 40% |
| - CIN2/3 | 79% |
| - Cancer | 82% |
| Cytology ≥ Pap3a2,[a] if no hrHPV infection is present | < 0.1% |
| Cytology ≥ Pap 3a2, if hrHPV infection is present | |
| - No CIN | 3% |
| - CIN1 | 5% |
| - CIN2/3 | 48% |
| - Cancer | 55% |

[a] *≥ Pap2 = abnormal cytology; ≥ Pap3a2 = HSIL.*

### A.2 Test characteristics

Characteristics of hrHPV screening tests are based on a population-based Dutch screening trial on self-collected samples versus clinician-collected smears.[11] The test characteristics of cytology are based on the results of meta-analyses of the performance of cytology after an HPV-positive test.[12]

### A.3 Adjustment in referral rates due to genotyping

In 2017, when the Dutch screening program for cervical cancer switched from primary cytology to primary HPV-testing with cytology triage, all HPV-positive women with cytological ASC-US (or higher) were referred for colposcopy. Since July 2022, the referral policy is based on HPV-16/18 genotyping in combination with the cytology result. Women with ASC-US/LSIL who test positive for HPV-16/18 are referred for colposcopy directly, while women who are hrHPV positive but negative for HPV-16/18 are referred only if repeat cytology is abnormal, otherwise they are reinvited at the next screening round. In order to adjust the model outcomes for this change in referral policy, we retrospectively calculated the referral rates of the first two rounds of the POBASCAM trial.[13]

The referral rate among HPV-positive women in the first two rounds under the 2017 policy without HPV-16/18 genotyping was 58.8%, while the referral rate under the 2022 policy was 51.9%.[13] The model-predicted rate of colposcopies and CIN diagnoses were adjusted for this policy change by the following genotype multiplier (*g*), which takes into account vaccination coverage (*c*) and type-specific vaccine efficacy (*ve*),

$$g = 1 - \left(1 - \frac{51.9}{58.9}\right)(1 - c) - (\left(1 - \frac{51.9}{58.9}\right)(1 - ve)\, c\,).$$

In the extreme cases where, *c=0*, i.e., no vaccination coverage, or *ve=0*, i.e., no vaccine efficacy, *g* would equal 1 – (1- 51.9/58.9) or 0.88, which means that the rate of colposcopies and CIN diagnoses predicted by the model would slightly decrease compared to no genotyping. For a vaccination coverage of 60% (observed in birth cohort 1999) and vaccine efficacy of 95%, *g* equals 0.95. We notice that for women who are vaccinated and protected, this policy change has no effect since these women will not test positive for HPV-16/18.

### A.4 Demographic and Health Parameters

#### A.4.1 Demography

Death due to other causes was simulated from the Dutch lifetable for the year 2000.[14]

#### A.4.2 Hysterectomy

Women who undergo hysterectomy are removed from the risk set for cervical cancer. Hysterectomy rates in the model for the Netherlands were based on the publication of Hanstede et al. 2012.[15] This publication uses data from 166,000 women who underwent a hysterectomy between 1995 and 2005. We counted the number of hysterectomies by age group assuming that the population age distribution did not change over this time period. Furthermore, we assumed that the age distribution of hysterectomies in the year 2005 is a good approximation to the age distribution of hysterectomies in 2023. Data on the size of the female population by age group in the year 2005 were retrieved from Statistics Netherlands (CBS).[16]

**Table S.5: Hysterectomy rates by age group.**

| Age (years) | Cumulative percentage of women with hysterectomy |
|---|---|
| 20 | 0% |
| 30 | 0.09% |
| 40 | 0.3% |
| 50 | 1.2% |
| 60 | 1.8% |

*Rates calculated from Hanstede et al. 2012* [15]

#### A.4.3 Survival from cervical cancer

Cancer survival is assigned at the time of diagnosis and is based on Dutch registry data.[8]

**Table S.6: Survival from diagnosis by cervical cancer stage.**

| | 1 year | 3 years | 5 years |
|---|---|---|---|
| FIGO1a | 0.99 | 0.95 | 0.93 |
| FIGO1b | 0.97 | 0.88 | 0.86 |
| FIGO2+ | 0.85 | 0.61 | 0.43 |

*Mean survival predicted by the model at 5 years (conditional on the predicted stage distribution at diagnosis) is 0.75 which compares to observed 0.74 in Dutch cancer registry data.*

### A.5 Vaccination coverage

Women born before 1997 were eligible for catch-up vaccination in 2009, if residing in the Netherlands at the time. Vaccination coverage in these catch-up cohorts is based on the 2011 RIVM report. Women born in or after 1997 received an invitation for routine HPV vaccination in the calendar year of their 13th birthday, with the option of free vaccination until age 18 through a delayed opt-in scheme. Vaccination coverage in these cohorts is based on the rates without age restriction, as reported in the 2020 RIVM report.

**Table S.7 Vaccination coverage in the Netherlands by birth cohort.**

| Birth Cohort | Year of vaccination | | | | | | |
|---|---|---|---|---|---|---|---|
| | 2009 | 2010 | 2011 | 2012 | 2013 | 2014 | 2015 |
| 1993 | 49% | 0 | 0 | 0 | 0 | 0 | 0 |
| 1994 | 52,5% | 0 | 0 | 0 | 0 | 0 | 0 |
| 1995 | 53,8% | 0 | 0 | 0 | 0 | 0 | 0 |
| 1996 | 54,2% | 0 | 0 | 0 | 0 | 0 | 0 |
| 1997 | 0 | 57,3% | 0 | 0 | 0 | 0 | 0 |
| 1998 | 0 | 0 | 58,8% | 0 | 0 | 0 | 0 |
| 1999 | 0 | 0 | 0 | 60,1% | 0 | 0 | 0 |
| 2000 | 0 | 0 | 0 | 0 | 61,3% | 0 | 0 |
| 2001 | 0 | 0 | 0 | 0 | 0 | 61,7% | 0 |
| 2002 | 0 | 0 | 0 | 0 | 0 | 0 | 57,1% |

### A.6 Screening attendance

From the age of 20 onward, i.e., before women enter the organized screening program, we assume an opportunistic uptake equal to 9.1% per five years, based on the observed rate of opportunistic screening attendance between age 20-30 in the Netherlands.[17] We model two types of attenders with respect to the organized screening program. The low attendance group, assumed to be 10% of the population, does not attend organized screening and has an opportunistic attendance rate that is based on the observed rate of opportunistic screening in the Netherlands in 2019, for the age groups included in the screening program, i.e. 30 to 65 years (8.4%). The high attendance group, assumed to be 90% of the population, is inclined to participate in organized screening, but has per-round probability of attending. For those women, attendance was reweighted such that the average attendance equaled the observed attendance (Table S.8).[18]

**Table S.8: Screening attendance in the Netherlands by five-year age group.**

| | 30-34 | 35-39 | 40-44 | 45-49 | 50-54 | 55-59 | 60-64 |
|---|---|---|---|---|---|---|---|
| **Attendance** | 65% | 71% | 74% | 74% | 78% | 78% | 77% |

*Based on the Dutch screening data, we assume compliance with repeat cytology after 12 months to be 81%. If a woman is referred to the gynecologist, we assume that the attendance is 90%. At age 65 attendance is assumed to be similar to that at age 60.*

#### A.7 CIN diagnosis and treatment costs

The number of diagnostic procedures were calculated from a population-based screening trial linked to the nationwide network and registry of histo- and cytopathology.[11,19] The CIN diagnosis costs included the number of indicative screening tests after colposcopy referral and before diagnosis, the number of follow-up screening tests after treatment (either cytology or HPV-testing), and the number of colposcopy-guided biopsies. Based on observed Dutch data, we assumed that all CIN3 cases and 65% of CIN2 cases were eventually treated and that repeat treatment was needed in 15% of the cases.[7,19,20] Unit costs for the indicative and follow-up screening tests were based on the subsidy regulation to the cervical cancer screening program determined by the Dutch Ministry of Welfare and Sports,[21] unit LLETZ (Large Loop Excision of the Transformation Zone) treatment costs were taken from the literature.[22] All costs were indexed to the year 2023.

#### A.8 HPV-FRAME Checklist

This study adheres to HPV-FRAME, a quality framework for the reporting of mathematical modelling evaluations of HPV-related cancer control. The checklist to be reported is separated into a core set of reporting measures (Table S.10) and a set specific for reporting on the outcomes of HPV vaccination in adolescent individuals (Table S.11).

**Table S.9: Calculation of CIN follow-up and treatment costs.**

| Cost category | Procedures per patient | Cost per medical procedure (€) | Cost per patient |
|---|---|---|---|
| **No CIN** | | | |
| Colposcopies | 2.04 | First: 191.7<br>Repeat: 158.8 | 357 |
| Biopsies | 1.37 | 70.1 | 96 |
| Co-tests | 0.67 | 60.6 | 41 |
| **Total** | | | **494** |
| **CIN 1** | | | |
| Colposcopies | 3.18 | First: 191.7<br>Repeat: 158.8 | 538 |
| Biopsies | 1.37 | 70.1 | 96 |
| Co-Tests | 1.81 | 60.6 | 110 |
| **Total** | | | **744** |
| **CIN 2** | | | |
| Colposcopies | 4.56 | First: 191.7<br>Repeat: 158.8 | 757 |
| Biopsies | 2.01 | 70.1 | 141 |
| Treatment (LLETZ) | 0.75 | 691 | 518 |
| Co-tests (cytology + HPV) after treatment | 2.55 | 60.6 | 155 |
| **Total** | | | **1571** |
| **CIN3** | | | |
| Colposcopies | 4.89 | First: 191.7<br>Repeat: 158.8 | 809 |
| Biopsies | 2.30 | 70.1 | 161 |
| Treatment (LLETZ) | 1.15 | 691 | 795 |
| Co-tests (cytology + HPV) after treatment | 2.59 | 60.6 | 157 |
| **Total** | | | **1922** |

### Table S.10: HPV-FRAME Checklist, Core Reporting Standard.

| **Core Reporting Standard** | | | |
|---|---|---|---|
| **Inputs** | **Reported by age?** | **Report by sex?** | **Comments** |
| Target population for intervention | Y | Y | Only vaccination in girls and women were considered. Age of routine and catch-up vaccination were reported, and screening of women between ages 30 and 65. |
| Sexual behavior | Y | Y | HPV incidence computed based on the HPV transmission model |
| Cohort examined for evaluation / time horizon | Y | Y | Multi birth cohort model representing Dutch population, followed from age 30 to end of life. |
| Quality of life assumptions | N | NA | Based on another Dutch publication, Jansen et al. [23] (Table 2); As a sensitivity analyses, we also used an alternative set of weights (Coupe et al. [10] and van Rosmalen et al. [24] |
| Calibration | Y | NA | Cervical cancer progression model calibration is described in sections A1.3. |
| Validation (where possible) | Y | NA | HPV prevalence, CIN detection and cervical cancer incidence predicted by the model were calibrated against observed data in the Netherlands. |
| Costs | N | NA | See Section A.7. All costs valued in Euros, and discounted at 3% according to Dutch 2024 guidelines. Costs also discounted at 3% as a sensitivity analysis based on WHO guidelines and at 4% based on 2016 Dutch guidelines. |

*Y= Yes, N=No, NA=Not Applicable.*

### Table S.11: HPV-FRAME Checklist, Reporting standard for HPV vaccination in adolescent individuals.

| Reporting standard for HPV vaccination in adolescent individuals | | | | |
|---|---|---|---|---|
| **Inputs** | **Reported** | **Reported by age?** | **Report by sex?** | **Comments** |
| Vaccine uptake | Y | Y | Y | Only vaccination in girls and women were considered. Uptake based on RIVM reports on vaccination coverage in routine and catch-up vaccination. |
| Vaccine efficacy | Y | NA | NA | Efficacy by HPV type was considered independent of age. |
| Vaccine cross-protection | Y | Y | NA | Level of cross-protection for HPV 31/33/45 was reported. |
| Duration vaccine protection and waning | Y | N | NA | Lifetime protection. |
| Vaccine and delivery costs | N | NA | NA | Vaccine and delivery costs are not considered because the intervention applies to screening of partly HPV-vaccinated cohorts. |
| Pre-vaccination disease burden (including population attributable fractions for HPV) | Y | Y | NA | Current HPV prevalence, CIN detection and cervical cancer incidence considered at baseline. |
| Duration of natural immunity | Y | N | NA | Natural immunity was independent of age. |
| **Outputs** | **Reported** | **Reported by age?** | **Report by sex?** | **Report as calibration or validation target? (Y/N)** |
| Absolute reductions in HPV infections, and/or warts, post-vaccination | N | N | N | HPV prevalence prior to vaccination was a calibration target. |
| Absolute reductions in CIN2+ post-vaccination | Y | N | N | CIN2+ detection was a calibration target and an output in evaluation of screening. |
| Absolute reductions in invasive cancer (cervical and other HPV cancers, as relevant) | Y, for cervical cancer | N | NA | Yes, **it** was a validation target for cancer progression model calibration. and an output in evaluation of screening. (See Figure A.2) |

[a] *Y= Yes, N=No, NA=Not Applicable, F= Female, ICER= Incremental Cost Effectiveness Ratio.*

### Supplementary Appendix B: Additional Model Results

#### B.1 Additional Model Results (base-case)

**Figure S.5: Cancer incidence with vaccination by birth cohort and current screening policy.**

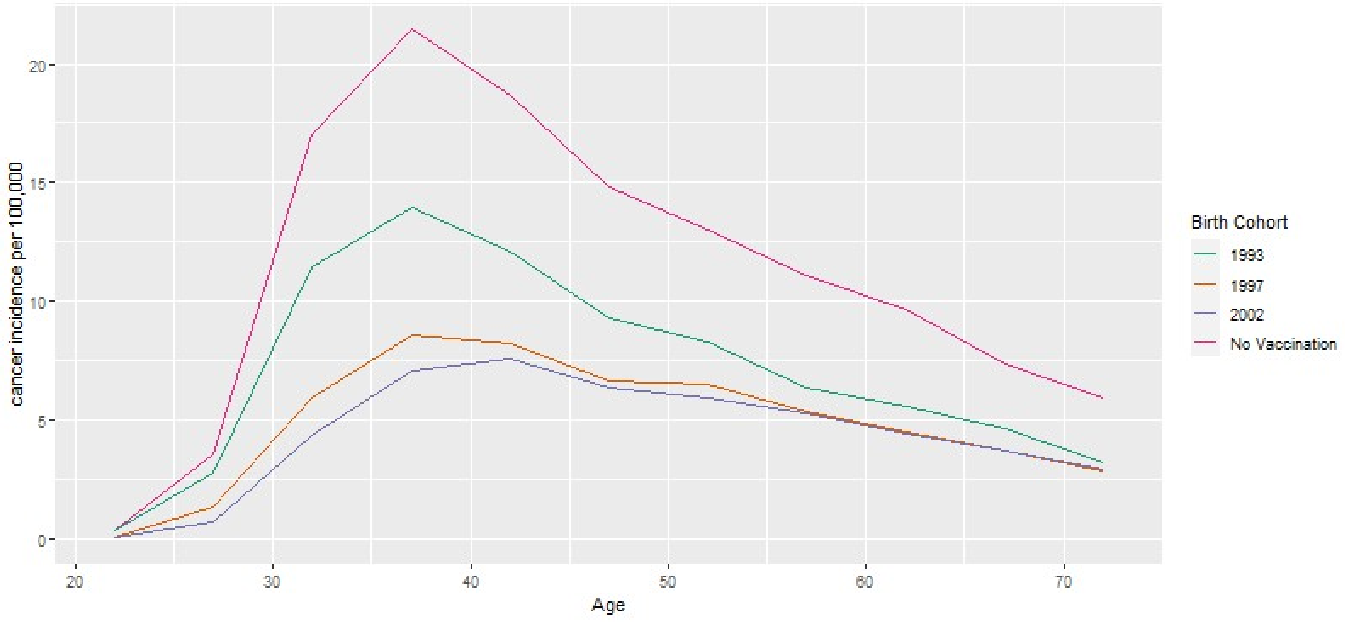

**Table S.12: Health and economic outcomes of cervical cancer screening in HPV-vaccinated cohorts.**

| Screening Policies [a] | Cancers (100,000 women) | Costs [b] (millions) | LY [b] (woman) | QALY [b] (woman) | NNS (CIN2+) | NNR (CIN2+) | NMB (millions) |
|---|---|---|---|---|---|---|---|
| *No Vaccination* | | | | | | | |
| NVS | 4172 | 43.8 | 32.5170 | 32.5073 | 0 | 0.0 | NA |
| B.NV | 633 | 27.7 | 32.6684 | 32.6618 | 64 | 2.0 | NA |
| *Vaccination (screening changes for all women)* | | | | | | | |
| NS | 2035 | 21.6 | 32.6083 | 32.6035 | 0 | 0.0 | NA |
| B | 294 | 19.6 | 32.6832 | 32.6786 | 130 | 2.5 | 0 |
| N30.1 | 420 | 17.0 | 32.6769 | 32.6733 | 117 | 2.1 | -8.1 |
| N30.2 | 424 | 16.4 | 32.6766 | 32.6732 | 116 | 2.1 | -7.7 |
| N30.3 | 569 | 16.1 | 32.6702 | 32.6670 | 108 | 1.87 | -19.8 |
| R35.1 | 304 | 18.0 | 32.6828 | 32.6786 | 118 | 2.4 | 1.65 |
| R35.2 | 392 | 15.9 | 32.6793 | 32.6756 | 94 | 2.2 | -2.3 |
| N35.1 | 473 | 16.8 | 32.6758 | 32.6721 | 105 | 2.0 | -10.3 |
| R | 313 | 17.5 | 32.6825 | 32.6784 | 113 | 2.4 | 1.8 |
| N40.1 | 390 | 18.1 | 32.6794 | 32.6753 | 122 | 2.2 | -5.2 |
| *Vaccination (screening only changes for vaccinated women)* | | | | | | | |
| N30.1 | 333 | 18.0 | 32.6815 | 32.6774 | 121 | 2.4 | -0.8 |
| N30.2 | 333 | 17.7 | 32.6814 | 32.6774 | 120 | 2.4 | -0.5 |
| N30.3 | 375 | 17.2 | 32.6795 | 32.6756 | 115 | 2.2 | -3.7 |
| R35.1 | 298 | 18.7 | 32.6830 | 32.6786 | 122 | 2.5 | 0.9 |
| R35.2 | 322 | 17.4 | 32.6821 | 32.6780 | 108 | 2.4 | 0.9 |
| N35.1 | 348 | 17.8 | 32.6811 | 32.6770 | 114 | 2.3 | -1.5 |
| R | 300 | 18.4 | 32.6830 | 32.6787 | 120 | 2.5 | 1.3 |
| N40.1 | 322 | 18.7 | 32.6821 | 32.6777 | 124 | 2.4 | -0.8 |

[a] *NVS denotes no vaccination and no screening. B denotes current screening policy. NS denotes no screening. NV denotes no vaccination. For details on each screening policy see Table 1.*

[b] *Costs are discounted at 3%, LYs and QALYs are discounted at 1.5%,*

**Table S.13: Additional undiscounted model outcomes for current screening and alternative screening policies in HPV-vaccinated cohorts.**

| Screening Policies [a] | Costs (millions) | LY (woman) | QALY (woman) | NNS (CIN3+) | NNR (CIN 3+) | Referrals | CIN leasions |
|---|---|---|---|---|---|---|---|
| *No Vaccination* | | | | | | | |
| B.NVS | 110.8 | 50.0745 | 50.0587 | 0 | 0.0 | 0 | 0 |
| B.NV | 45.9 | 50.3674 | 50.3590 | 95 | 3.0 | 12198 | 7132 |
| *Vaccination (screening changes for all women)* | | | | | | | |
| B.NS | 54.2 | 50.2497 | 50.2420 | 0 | 0.0 | 0 | 0 |
| B | 32.2 | 50.3939 | 50.3881 | 211 | 4.1 | 7798 | 4070 |
| N30.1 | 30.2 | 50.3831 | 50.3783 | 177 | 3.2 | 6089 | 3414 |
| N30.2 | 29.8 | 50.3825 | 50.3780 | 173 | 3.1 | 5912 | 3366 |
| N30.3 | 30.4 | 50.3704 | 50.3660 | 151 | 2.5 | 4648 | 2794 |
| R35.1 | 30.1 | 50.3932 | 50.3879 | 188 | 3.9 | 7388 | 3948 |
| R35.2 | 27.2 | 50.3866 | 50.3819 | 142 | 3.3 | 6153 | 3444 |
| N35 | 29.3 | 50.3801 | 50.3753 | 154 | 3.0 | 5532 | 3125 |
| R | 29.5 | 50.3926 | 50.3875 | 180 | 3.8 | 7193 | 3882 |
| N40.1 | 31.1 | 50.3866 | 50.3813 | 186 | 3.4 | 6467 | 3526 |
| *Vaccination (screening only changes for vaccinated women)* | | | | | | | |
| N30.1 | 30.6 | 50.3909 | 50.3857 | 191 | 3.7 | 7077 | 3812 |
| N30.2 | 30.4 | 50.3907 | 50.3856 | 189 | 3.7 | 6999 | 3791 |
| N30.3 | 30.0 | 50.3871 | 50.3821 | 177 | 3.4 | 6480 | 3573 |
| R35.1 | 31.0 | 50.3935 | 50.3880 | 197 | 4.0 | 7621 | 4020 |
| R35.2 | 29.0 | 50.3919 | 50.3867 | 171 | 3.8 | 7097 | 3823 |
| N35.1 | 29.8 | 50.3900 | 50.3847 | 177 | 3.6 | 6850 | 3704 |
| R | 30.6 | 50.3935 | 50.3881 | 193 | 4.0 | 7542 | 3995 |
| N40.1 | 31.2 | 50.3917 | 50.3862 | 196 | 3.8 | 7236 | 3853 |

[a] *NVS denotes no vaccination and no screening. B denotes current screening policy. NS denotes no screening. NV denotes no vaccination. For details on each screening policy see Table 1.*

### B.2 Key Scenario and Sensitivity Analyses

**Figure S.6: Costs and health gains of reduced screening in partly HPV-vaccinated cohorts under 100% screen attendance.**

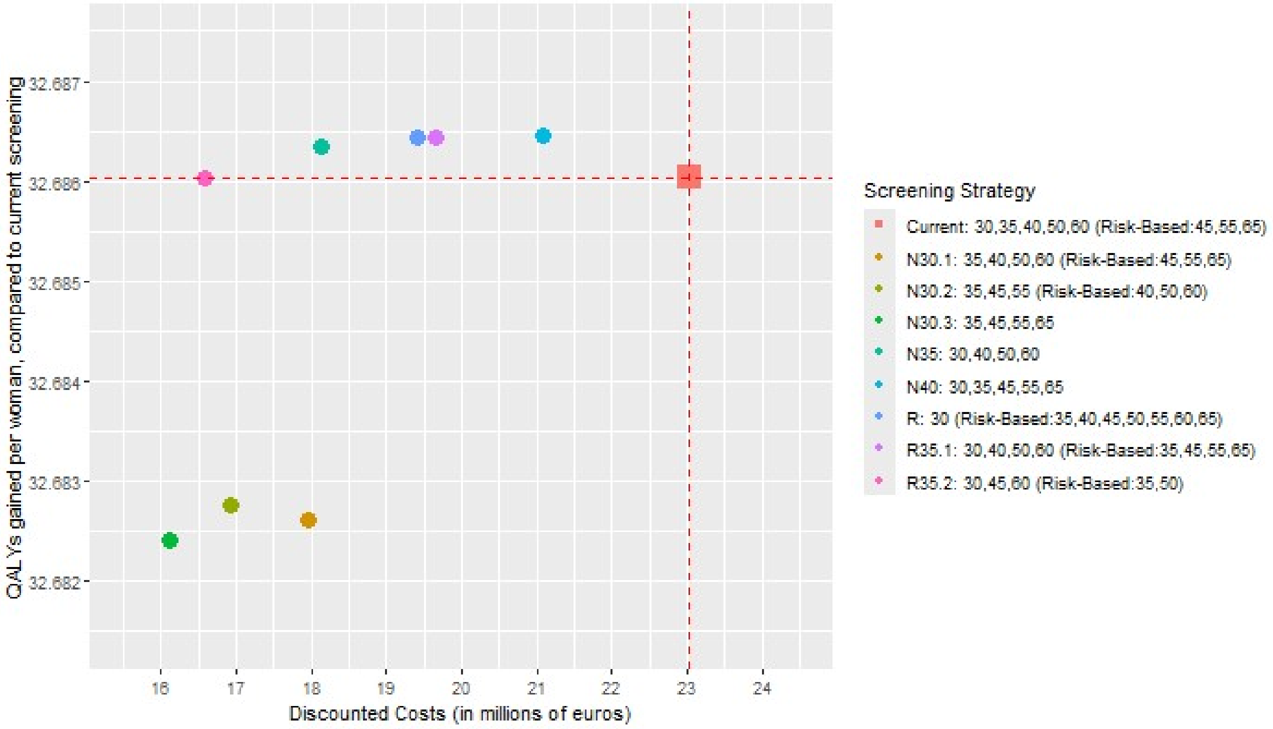

*Discounted costs at 3% per 100,000 women. In the risk-based rounds, only women who tested HPV-positive in the previous round, or who did not attend the previous round, are invited.*

**Figure S.7: Net monetary benefit and cancer incidence relative to current policy without vaccination under 100% screen attendance.**

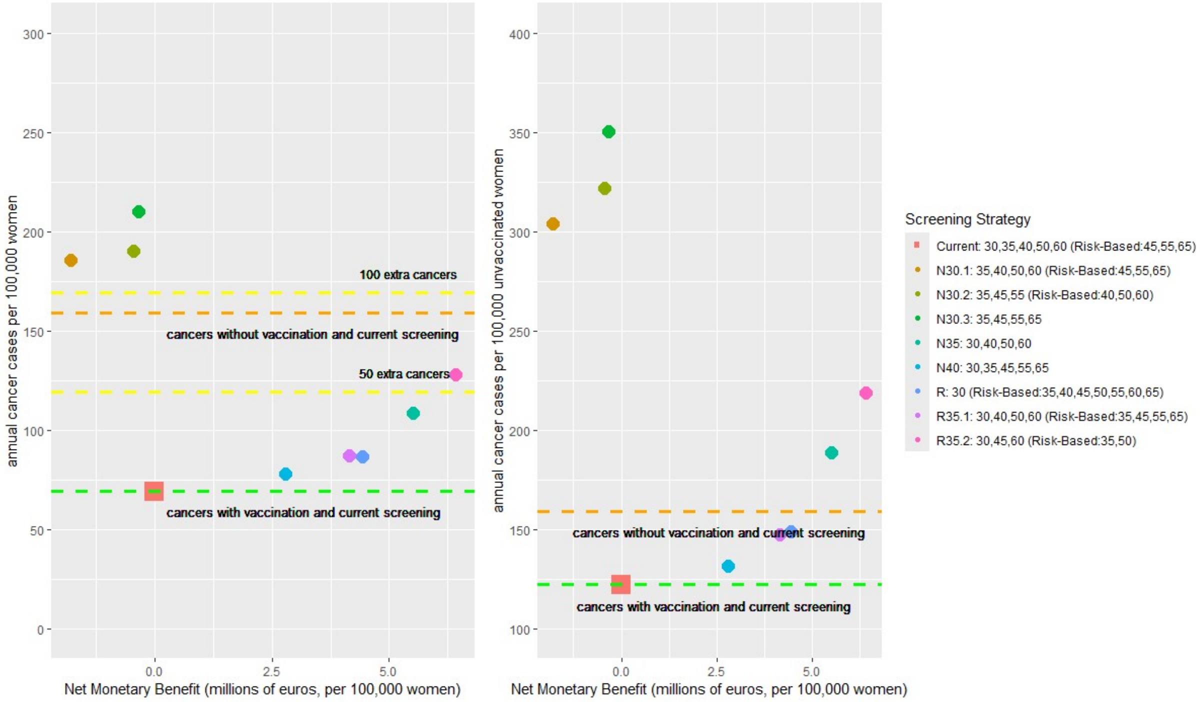

*Costs are discounted at 3% and QALYs are discounted at 1.5%. In the risk-based rounds, only women who tested HPV-positive in the previous round, or who did not attend the previous round, are invited.*

**Figure S.8: Costs and health gains of reduced screening in partly HPV-vaccinated cohorts under alternative QALY weights.**

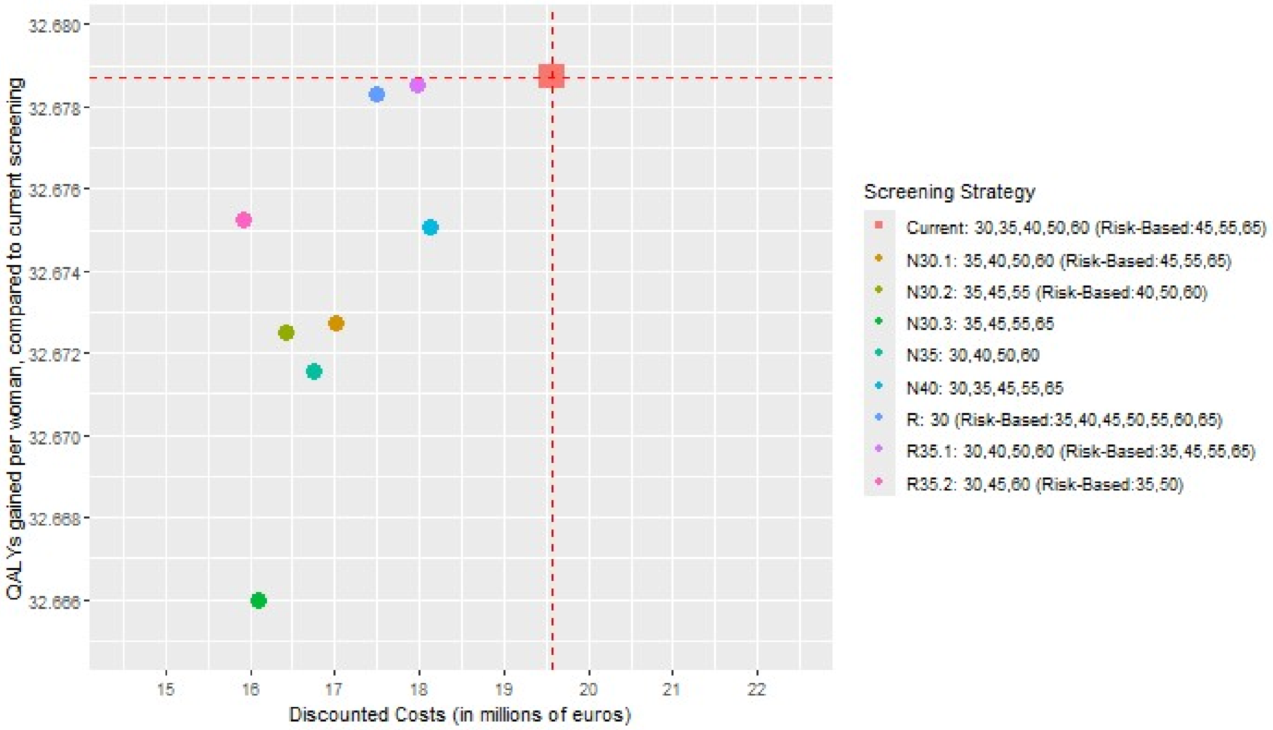

*Discounted costs at 3% per 100,000 women. In the risk-based rounds, only women who tested HPV-positive in the previous round, or who did not attend the previous round, are invited.*

**Figure S.9: Net monetary benefit and cancer incidence relative to current policy without vaccination under alternative QALY weights.**

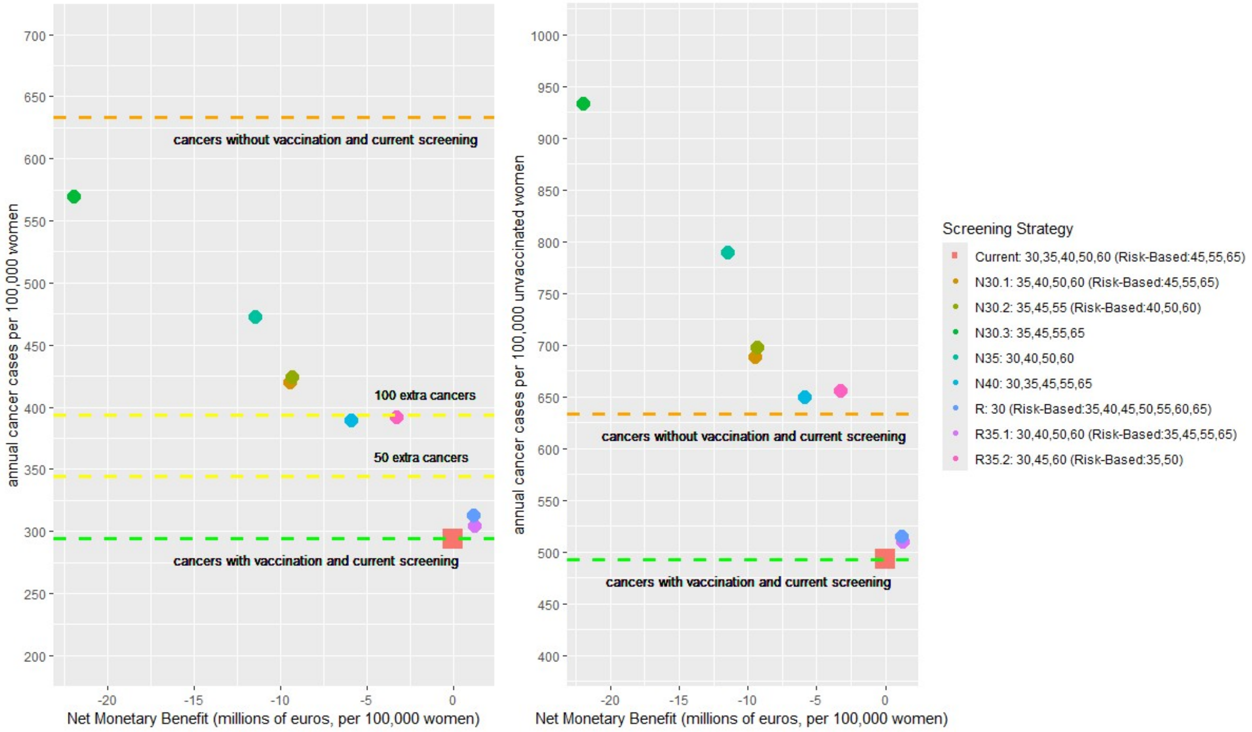

*Costs are discounted at 3% and QALYs are discounted at 1.5%. In the risk-based rounds, only women who tested HPV-positive in the previous round, or who did not attend the previous round, are invited.*

**Figure S.10: Costs and health gains of reduced screening in partly HPV-vaccinated cohorts under conservative vaccine efficacy.**

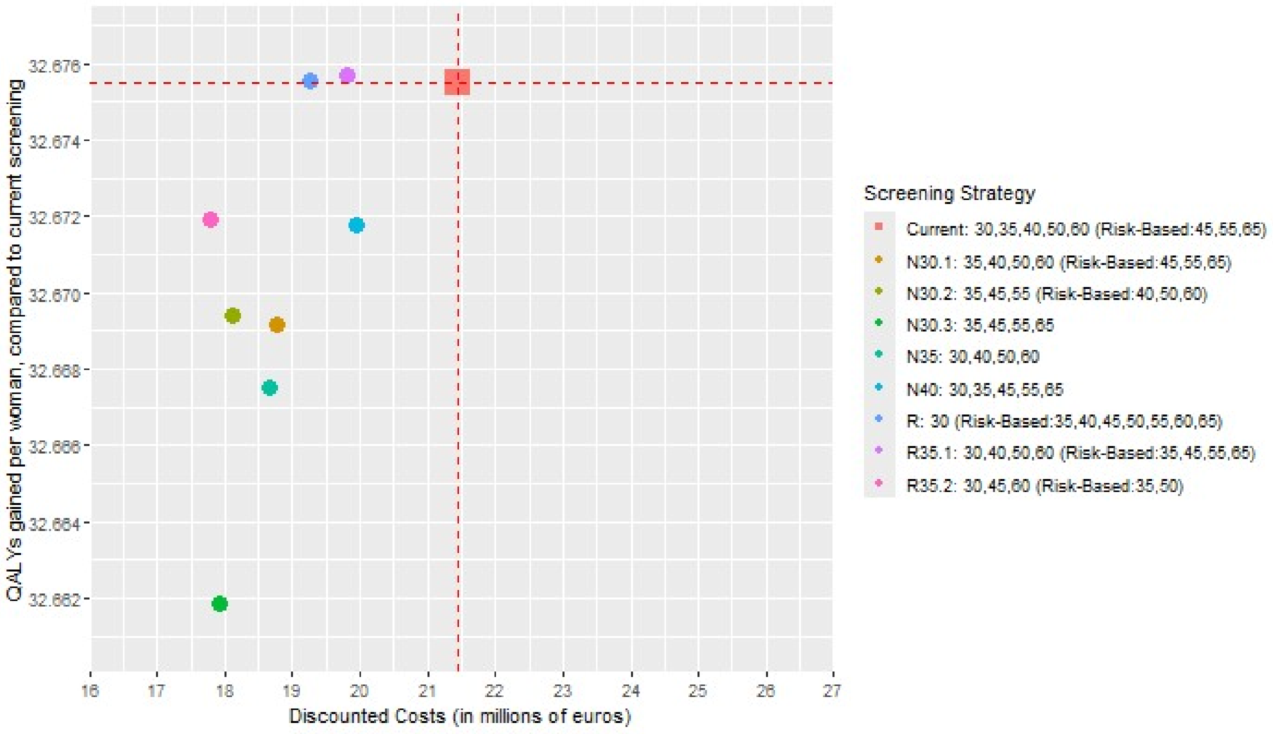

*Discounted costs at 3% per 100,000 women. In the risk-based rounds, only women who tested HPV-positive in the previous round, or who did not attend the previous round, are invited.*

**Figure S.11: Net monetary benefit and cancer incidence relative to current policy without vaccination under conservative vaccine efficacy.**

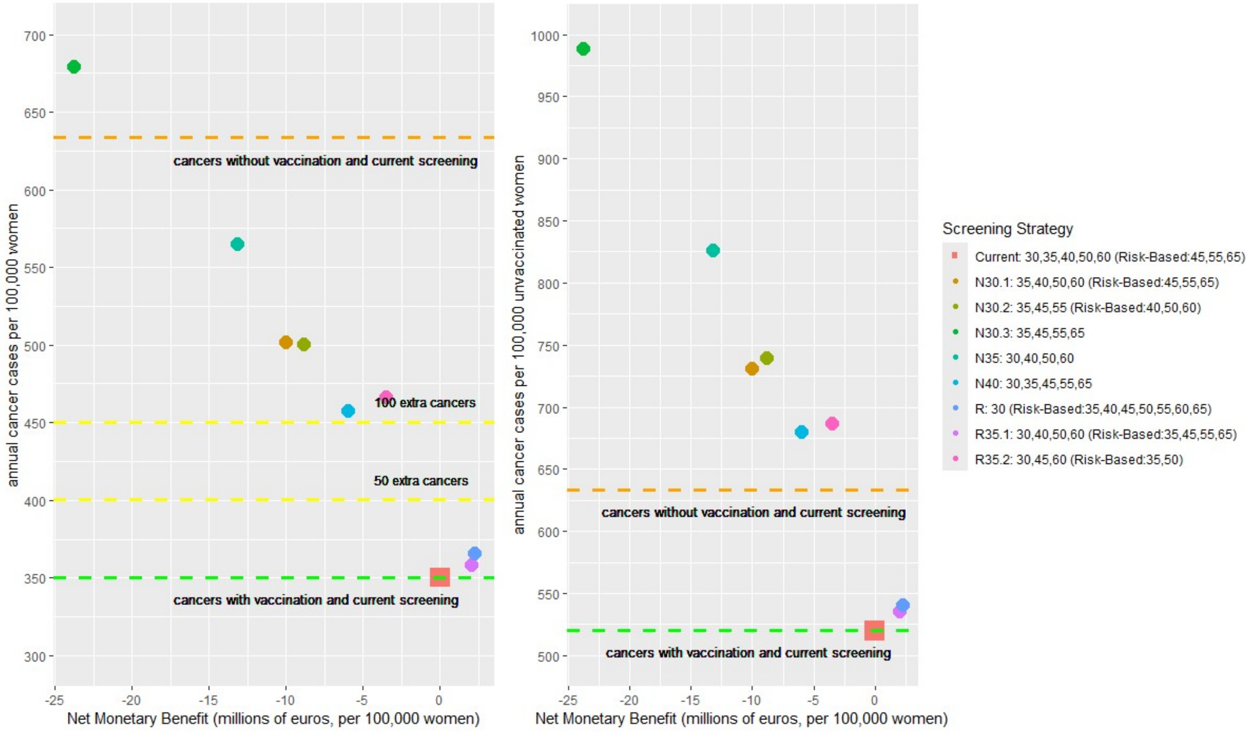

*Costs are discounted at 3% and QALYs are discounted at 1.5%. In the risk-based rounds, only women who tested HPV-positive in the previous round, or who did not attend the previous round, are invited.*
